# Cannabis Co-Use and Endocannabinoid System Modulation in Tobacco Use Disorder: A Translational Systematic Review and Meta-Analysis

**DOI:** 10.64898/2026.02.12.26346166

**Authors:** Gabriel P. A. Costa, Oscar Gómez, Lucas R. da Rocha, Mayte A. Cerezo-Matias, Melissa C. Funaro, Deniz Bagdas, Mehmet Sofuoglu, Joao P. De Aquino

**Author notes:** **Corresponding Authors:** Joao P. De Aquino, M.D. Department of Psychiatry Yale School of Medicine 34 Park St, 333B, New Haven, CT 06519 Gabriel P. A. Costa, M.D. Department of Psychiatry Yale School of Medicine 34 Park St, 332, New Haven, CT 06519. These authors have contributed equally to this work and share senior authorship (Mehmet Sofuoglu, Joao P. De Aquino).

## Abstract

Tobacco use disorder (TUD) remains a leading cause of preventable mortality, and existing pharmacotherapies yield 12-month abstinence rates below 30%. As cannabis legalization expands, approximately 18-22% of people who use tobacco report concurrent cannabis use, yet the impact of co-use on cessation outcomes and the therapeutic potential of endocannabinoid system (ECS) modulation remain unclear. We conducted a translational systematic review and meta-analysis following PRISMA 2020 guidelines, searching Ovid MEDLINE, Embase, APA PsycInfo, and Web of Science through January 2026 (PROSPERO: CRD420250652724). Three study categories were eligible: observational studies of cannabis co-use and cessation outcomes; preclinical studies of cannabinoid modulators on nicotine-related behaviors; and human experimental studies of ECS-targeted interventions. Of 4,869 records screened, 52 studies met inclusion criteria. Meta-analysis of 18 observational studies (N=229,630) revealed that cannabis use was associated with 35% lower odds of achieving tobacco cessation (OR=0.65; 95% CI: 0.55-0.78; p<0.0001; I²=88.1%). Preclinical evidence (15 studies) demonstrated that CB1 receptor antagonists robustly reduced nicotine self-administration and reinstatement, while cannabidiol (CBD) attenuated both nicotine intake and withdrawal without affecting food reinforcement. Clinical translation of CB1 receptor inverse agonists failed due to psychiatric adverse effects, but CBD showed promise by reducing cigarette consumption by 40%, reversing attentional bias to smoking cues, and alleviating withdrawal severity. These findings distinguish naturalistic cannabis exposure from potentially beneficial targeted ECS modulation, and support CBD as a promising candidate for adequately powered tobacco cessation trials.

## 1. Introduction

Tobacco use disorder (TUD) is a leading cause of preventable mortality globally, accounting for more than 7 million deaths annually and imposing a substantial burden through cardiovascular disease, malignancy, and respiratory illness^1,2^. In the U.S., approximately 28.3 million adults currently use tobacco or electronic cigarettes (e-cigarettes), and despite decades of public health efforts, the majority of quit attempts fail; fewer than 10% of smokers achieve long-term abstinence without pharmacological support^3,4^. While FDA-approved pharmacological treatments, including nicotine replacement therapy (NRT), bupropion, and varenicline, increase cessation rates 2- to 3-fold compared to placebo^3,5^, many individuals do not benefit from these treatments. This therapeutic ceiling, combined with the limited range of pharmacological mechanisms targeted by available pharmacotherapies, underscores the need to identify novel therapeutic approaches for TUD.

Against the backdrop of limited treatments for TUD, the rapid expansion of cannabis legalization across North America and Europe has introduced the rising prevalence of cannabis-tobacco co-use. National survey data indicate that approximately 18-22% of people who currently use tobacco report concurrent cannabis use^6–8^. Cannabis and tobacco are frequently co-administered within the same product (e.g., blunts, spliffs), a practice that creates pharmacological and behavioral interactions that potentially reinforce the use of both substances^9–11^. Epidemiological evidence suggests that people who co-use tobacco and cannabis, particularly people who use cannabis daily, may exhibit greater nicotine dependence severity, increased frequency of failed quit attempts, and elevated relapse vulnerability, compared to people who use tobacco only. However, this association may not extend to people who use cannabis non-daily^9,11–13^. As cannabis access continues to expand, understanding whether and how co-use impacts tobacco and nicotine product cessation outcomes has become a clinical priority. Throughout this review, “cessation” refers to abstinence from combustible cigarettes, e-cigarettes, or both, as defined by the individual study.

### 1.1. Crosstalk Between Endocannabinoid and Nicotinic Cholinergic Pathways

The extensive crosstalk between the endocannabinoid system (ECS) and the nicotinic cholinergic receptors (nAChR) underscores the therapeutic potential of targeting the ECS for TUD. The endocannabinoid system (ECS) comprises cannabinoid receptors (primarily CB1 and CB2), endogenous ligands including anandamide (AEA) and 2-arachidonoylglycerol (2-AG)^14^.

The signaling activity of AEA and 2-AG is terminated primarily by fatty acid amide hydrolase (FAAH) and monoacylglycerol lipase (MAGL), respectively^15^. CB1 receptors (CB1R)^16^ are densely expressed throughout the brain reward-related circuitry, including the ventral tegmental area (VTA), nucleus accumbens (NAc), prefrontal cortex, and amygdala, where they serve as retrograde modulators of glutamatergic and GABAergic transmission^17,18^.

The nAChR are ligand-gated ion channels that are pentameric assemblies of 12 distinct subunits: α2-α10 and β2-β4^19,20^. In the brain, the most prevalent subtypes are heteromeric α4β2* *(* denotes possible inclusion of additional subunits)* and homomeric α7 nAChR. The activation of nAChR facilitates the release of key neurotransmitters, including dopamine, acetylcholine, GABA, and glutamate. Dopamine release in the NAc, central to the reinforcing effects of nicotine, is mediated primarily by α4β2* nAChR. The NAc also integrates GABAergic and glutamatergic inputs that inhibit and stimulate nicotine-induced DA release, respectively.

The nAChR subtypes that regulate GABA release (primarily α4β2*) desensitize more rapidly than those that regulate glutamate release (primarily α7)^21,22^. With chronic nicotine exposure, this relative deficiency of inhibitory GABA compared to excitatory glutamate represents a key mechanism facilitating dopamine release and the development of nicotine dependence.

Convergent evidence supports bidirectional modulation between ECS and nAChR. In rodent models, nicotine self-administration increases the release of AEA and 2AG from post- synaptic neurons in the NAc^23^. By activating presynaptic CB1R via retrograde signaling, these endocannabinoids reduce the release of both GABA and glutamate within NAc^24^. This reduced GABA release is suggested to amplify dopamine signaling by removing the inhibitory brake^25,26^. Critically, CB1R is essential in mediating the reinforcing effects of nicotine, as CB1R antagonists effectively block these reinforcing effects. Chronic nicotine exposure further alters endocannabinoid signaling^27^, with preliminary evidence suggesting that people who co-use cannabis and tobacco have higher FAAH activity and lower AEA levels across multiple brain regions, compared to those who exclusively use either substance^28^. Reduced AEA levels are associated with sleep disturbance, anxiety, and anhedonia, and greater sensitivity to physical discomfort and pain (e.g., hyperalgesia), and may contribute to increased withdrawal severity and lower cessation success among co-users^29,30^. Evidence from positron emission tomography (PET) imaging studies in humans also indicates that chronic nicotine exposure is associated with lower density of CB1R^31^, likely reflecting ECS adaptations induced by long-term nicotine use. Collectively, these findings underscore the ECS as a promising target for novel tobacco cessation treatments, particularly among individuals who co-use cannabis and tobacco.

This neurobiological intersection between the ECS and nAChR raises two complementary questions: First, if exogenous cannabinoid exposure engages the same circuitry that modulates nicotine reinforcement, does co-use of tobacco and cannabis alter tobacco cessation outcomes? Second, can pharmacological interventions targeting specific ECS components, distinct from uncontrolled cannabis exposure, provide therapeutic benefit for TUD?

To address these questions, we conducted a translational systematic review and meta-analysis. We quantitatively synthesized observational evidence on cannabis and tobacco co-use and cessation outcomes and systematically evaluated preclinical and human experimental studies of ECS modulators, including CB1R antagonists, endocannabinoid enzyme and transport inhibitors, and CBD, as potential therapeutic targets. In doing so, we sought to (1) quantify the clinical impact of cannabis co-use on cessation success, (2) identify ECS mechanisms with therapeutic promise, (3) elucidate translational barriers, and (4) clarify the variables determining the distinction between naturalistic cannabinoid exposure and targeted pharmacotherapy.

## 2. Methods

This systematic review and meta-analysis is reported in accordance with the Preferred Reporting Items for Systematic Reviews and Meta-Analyses (PRISMA) 2020 guidelines^32^. The protocol was prospectively registered with PROSPERO CRD420250652724.

### 2.1 Search Strategy

We searched Ovid MEDLINE, Embase, APA PsycInfo, and Web of Science from inception through December 2024, with a final search update conducted in January 2026. The search strategy combined terms related to the ECS (cannabinoid, endocannabinoid, CB1, CB2, anandamide, 2-AG, CBD, tetrahydrocannabinol, rimonabant, FAAH) with nicotine/tobacco-related terms (nicotine, tobacco, smoking, cigarette, cessation, withdrawal). Reference lists of eligible studies and relevant systematic reviews were manually screened. Complete search strategies are provided in **Supplementary eMethods**.

### 2.2 Eligibility Criteria

Given the review’s dual objectives, (1) evaluating whether naturalistic cannabis co-use is associated with tobacco cessation outcomes and (2) assessing whether targeted ECS modulation offers therapeutic potential for TUD, three study categories were eligible.

First, observational studies examining the association between cannabis use (exposure) and tobacco or nicotine product abstinence or relapse among individuals receiving any cessation intervention. Studies were required to report odds ratios, relative risks, or sufficient data permitting their calculation.

Second, preclinical studies comprising controlled experiments using animal models examining effects of cannabinoid system modulators on nicotine-consumption, including self-administration, reinstatement of nicotine use, and conditioned place preference.

Third, human experimental studies including randomized controlled trials, crossover designs, and laboratory paradigms evaluating cannabinoid interventions for tobacco-related outcomes (consumption, craving, withdrawal, cue reactivity, cessation) or examining mechanistic interactions between cannabis and tobacco.

### 2.3 Data Extraction and Synthesis

Two reviewers (G.C. and O.G.) independently screened titles/abstracts and full texts, with discrepancies resolved by consensus discussion. Data extraction employed standardized forms tailored to each study category. For observational studies, we extracted odds ratios (or calculated them from reported data), confidence intervals, sample sizes, cannabis use definitions, cessation outcomes, and adjustment variables.

Only the observational studies examining cannabis co-use and tobacco cessation outcomes were considered meta-analyzable, as they reported a common effect measure (odds ratios for cessation) with sufficient methodological homogeneity for quantitative pooling.

Quantitative synthesis of preclinical and human experimental studies was precluded by the heterogeneity in species, compounds, dosing regimens, outcome measures, and study designs.

For the meta-analyzable observational studies, we employed inverse-variance random-effects models with restricted maximum likelihood estimation of between-study variance (τ²).

Heterogeneity was assessed using Cochran’s Q statistic, τ², and I². R version 4.5.2 was used for all analyses. For preclinical and human experimental studies, synthesis was narratively organized by cannabinoid mechanism given heterogeneity in designs and outcomes.

### 2.4 Risk of Bias Assessment

We assessed risk of bias in randomized trials using the Cochrane Risk of Bias 2 (RoB 2) tool and observational studies using the Risk of Bias in Non-randomized Studies of Interventions (ROBINS-I) tool. No formal quality assessment was conducted for preclinical studies due to lack of widely accepted tools for animal studies.

## 3. Results

Database searches identified 4,869 records after deduplication. Following title and abstract screening and a full-text review of 216 articles, 52 studies met the inclusion criteria: 18 observational studies examining cannabis as a predictor of cessation, 15 preclinical studies evaluating cannabinoid modulators, and 19 human experimental studies (**Figure 2**).

**Figure 1.**
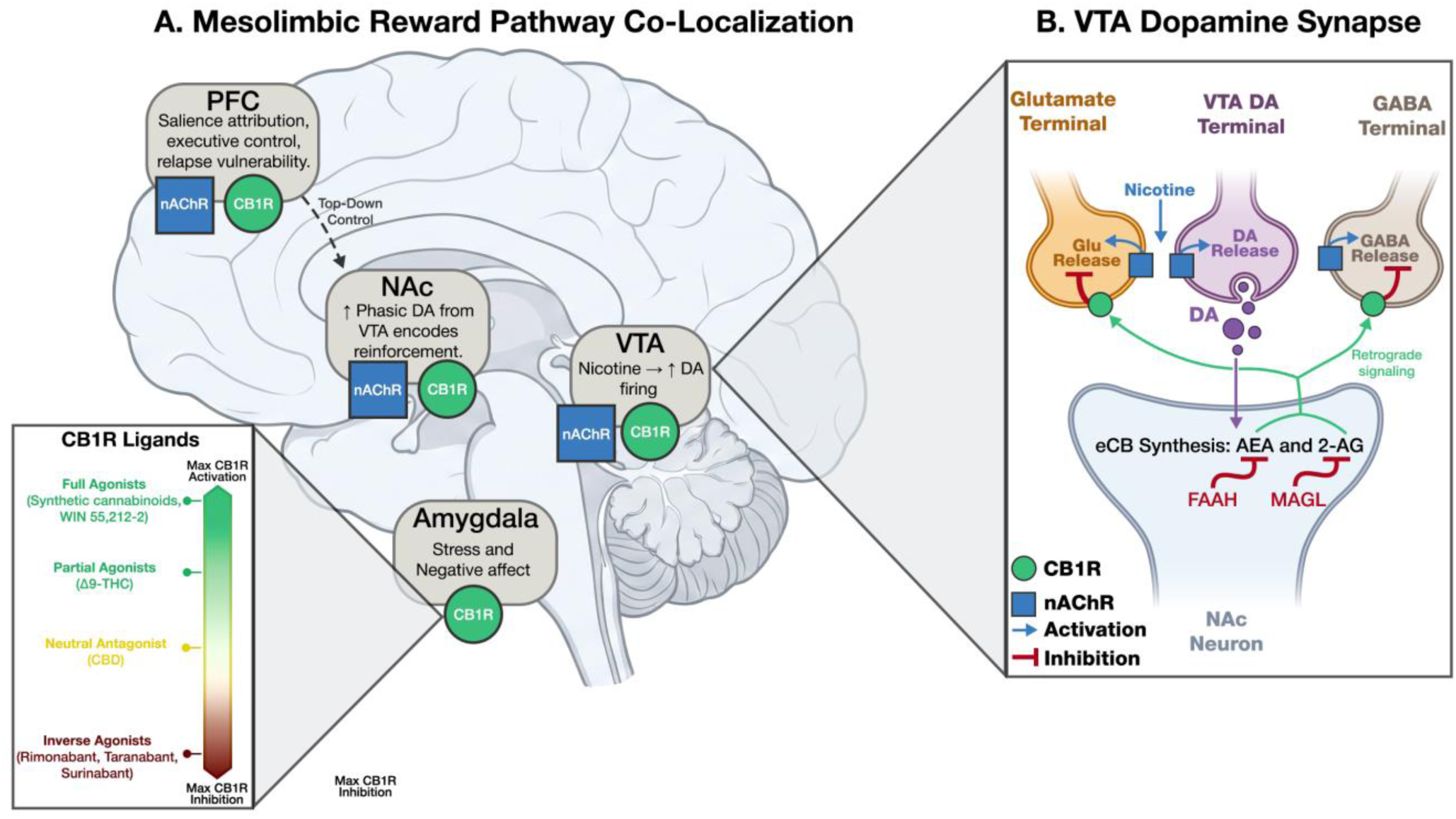
Endocannabinoid–Nicotinic Convergence in Mesolimbic Reward Circuitry. **(A)** Schematic highlighting co-localization of nicotinic acetylcholine receptors (nAChRs) and cannabinoid type-1 receptors (CB1R) across key nodes implicated in tobacco reinforcement and relapse vulnerability: prefrontal cortex (PFC; salience attribution/executive control), ventral tegmental area (VTA; nicotine-driven dopaminergic firing), nucleus accumbens (NAc; phasic dopamine [DA] encoding reinforcement), and amygdala (stress/negative affect). The CB1R ligand spectrum (inset) illustrates how different compounds engage CB1R with varying directionality and magnitude, from full agonists (e.g., WIN 55,212-2) and partial agonists (Δ9-THC) that activate the receptor, to negative allosteric modulators (CBD) that attenuate receptor signaling without suppressing constitutive activity, to inverse agonists (rimonabant, taranabant, surinabant) that suppress both agonist-driven and constitutive CB1R signaling. **(B)** VTA dopamine synapse model illustrating nicotine activation of α4β2* nAChRs, nicotine-evoked DA signaling, and activity-dependent synthesis of endocannabinoids (anandamide [AEA] and 2-arachidonoylglycerol [2-AG]). Endocannabinoids act as retrograde signals to activate presynaptic CB1R, suppressing GABA release and disinhibiting DA neurons. Endocannabinoid tone is terminated by fatty acid amide hydrolase (FAAH; AEA) and monoacylglycerol lipase (MAGL; 2-AG). Arrows indicate facilitation/activation; blunt lines indicate inhibition.

**Figure 2.**
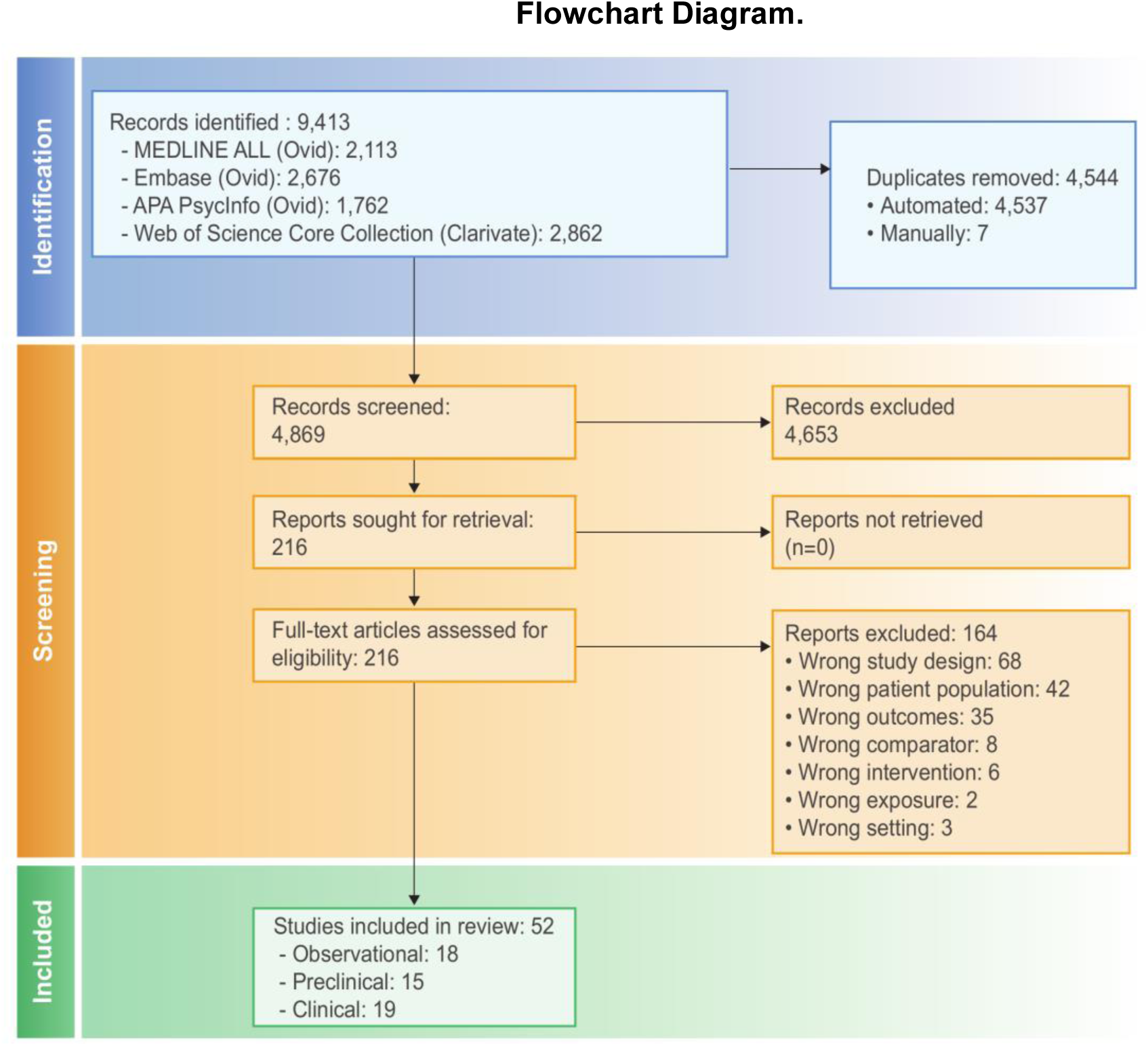
Preferred Reporting Items for Systematic Reviews and Meta-Analyses (PRISMA) Flowchart Diagram. Flow diagram illustrating the study selection process. A total of 9,413 records were identified across four databases. After removing 4,544 duplicates, 4,869 records were screened by title and abstract, and 216 full-text articles were assessed for eligibility. 52 studies met inclusion criteria: 18 observational, 15 preclinical, and 19 clinical.

The results are organized to first examine the impact of co-use on cessation (observational evidence), followed by an exploration of the therapeutic potential of cannabinoid interventions (preclinical and human experimental evidence).

### 3.1 Cannabis Co-Use and Tobacco Cessation: Observational Evidence

Eighteen observational studies contributing 20 independent estimates (total N=229,630) examined whether cannabis use predicted tobacco cessation outcomes among individuals receiving cessation interventions. Follow-up ranged from end-of-treatment to 12 months, with 6-month assessment most common.

Meta-analysis yielded a pooled odds ratio of 0.65 (95% CI: 0.55-0.78; p<0.0001), indicating that people who use cannabis have 35% lower odds of achieving cessation compared to people who do not use cannabis (**Figure 3**). Between-study heterogeneity was high (I²=88.1%; 95% CI: 83.1-91.7%; τ²=0.10; Cochran’s Q=160.24, df=19, p<0.0001), reflecting substantial variability in effect magnitude. Visual inspection of the funnel plot did not reveal marked asymmetry (**Supplement Figure S1**) and neither Egger’s regression test (p = 0.071) nor Begg’s rank correlation test (p = 0.948) indicated significant publication bias.

**Figure 3.**
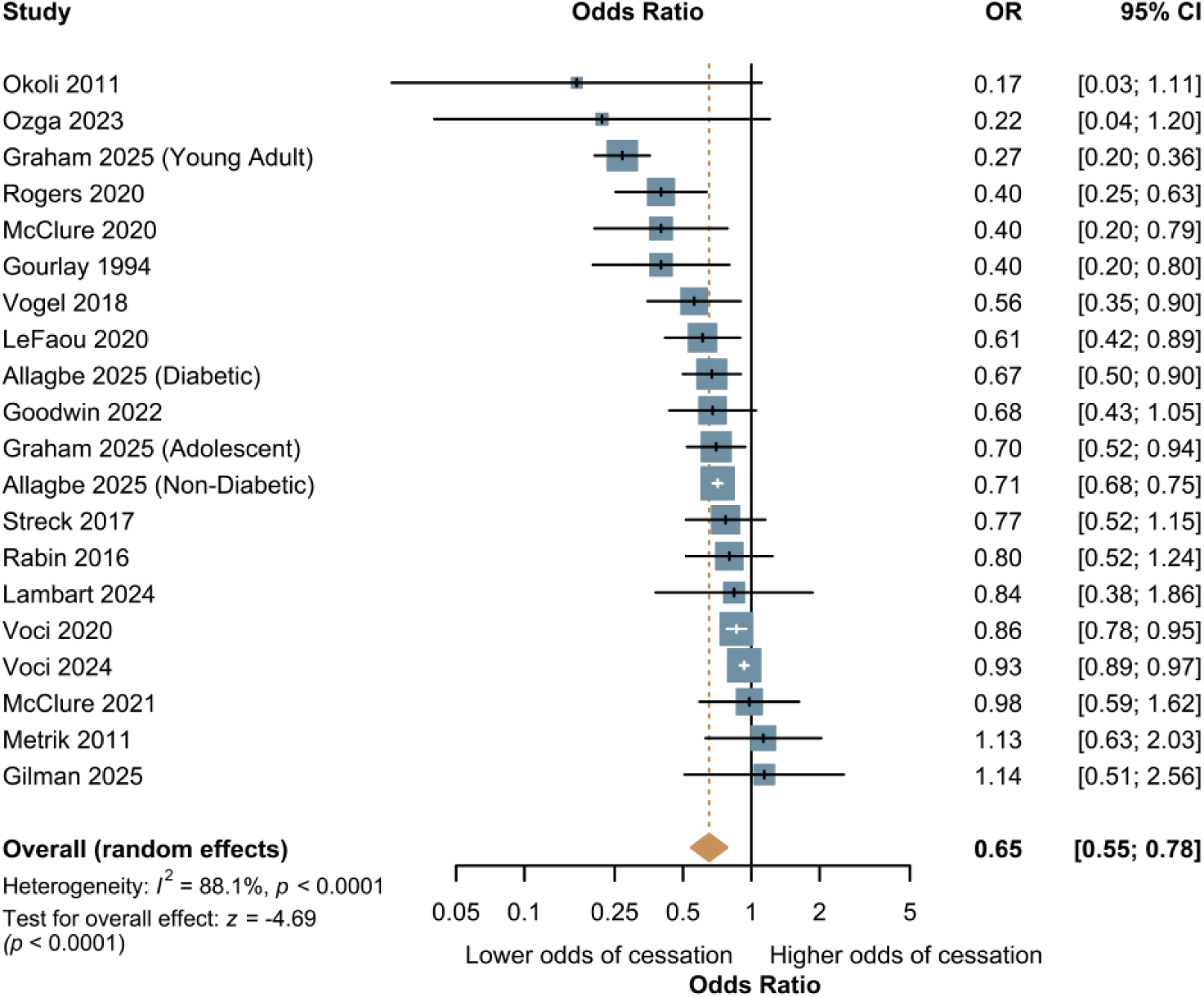
Forest Plot of the Association Between Cannabis Use and Tobacco Cessation Outcomes in Observational Studies. Random-effects meta-analysis of 18 observational studies (20 independent estimates; N=229,630) examining the odds of tobacco cessation among people who use cannabis compared to people who do not use cannabis. The pooled odds ratio was 0.65 (95% CI: 0.55-0.78; p<0.0001), indicating 35% lower odds of cessation among people who use cannabis. Between-study heterogeneity was high (I²=88.1%; p<0.0001). Individual study estimates are represented by squares (proportional to study weight), with horizontal lines indicating 95% confidence intervals. The diamond represents the pooled effect estimate.

### 3.2 Endocannabinoid System Modulation of Nicotine-Related Behaviors: Preclinical Evidence

Fifteen preclinical studies examined ECS modulation of nicotine-related behaviors using rodent and primate models across 17 experiments (**Table 1**). Paradigms included intravenous nicotine self-administration under fixed-ratio and progressive-ratio schedules, reinstatement models (cue-induced, nicotine-primed, contextual), conditioned place preference, and withdrawal assessment. Interventions were categorized by mechanism: CB1R antagonists/inverse agonists (n=6), CB1R agonists (n=2), CBD (n=1), and other endocannabinoid modulators (n=6).

**Table 1.**
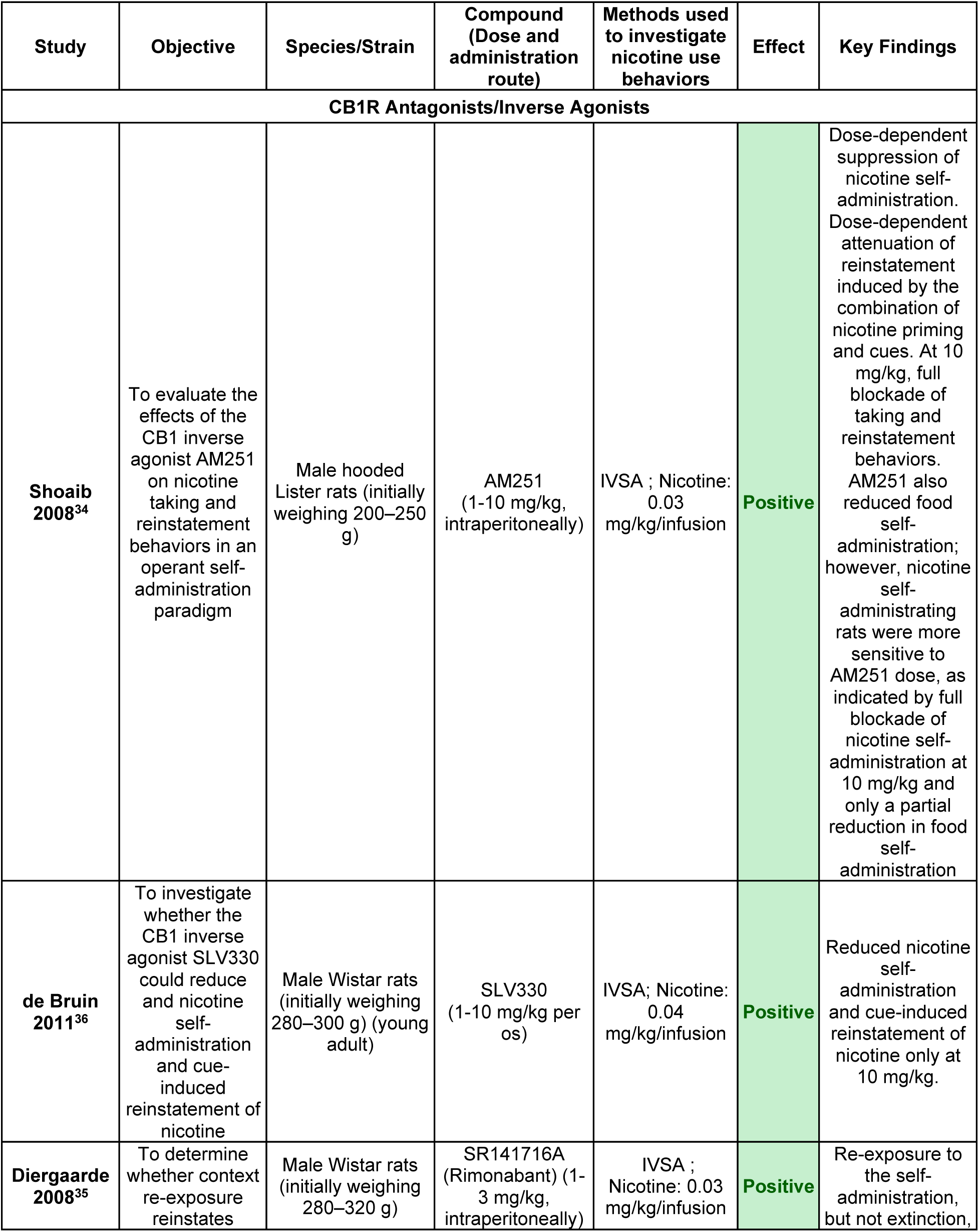
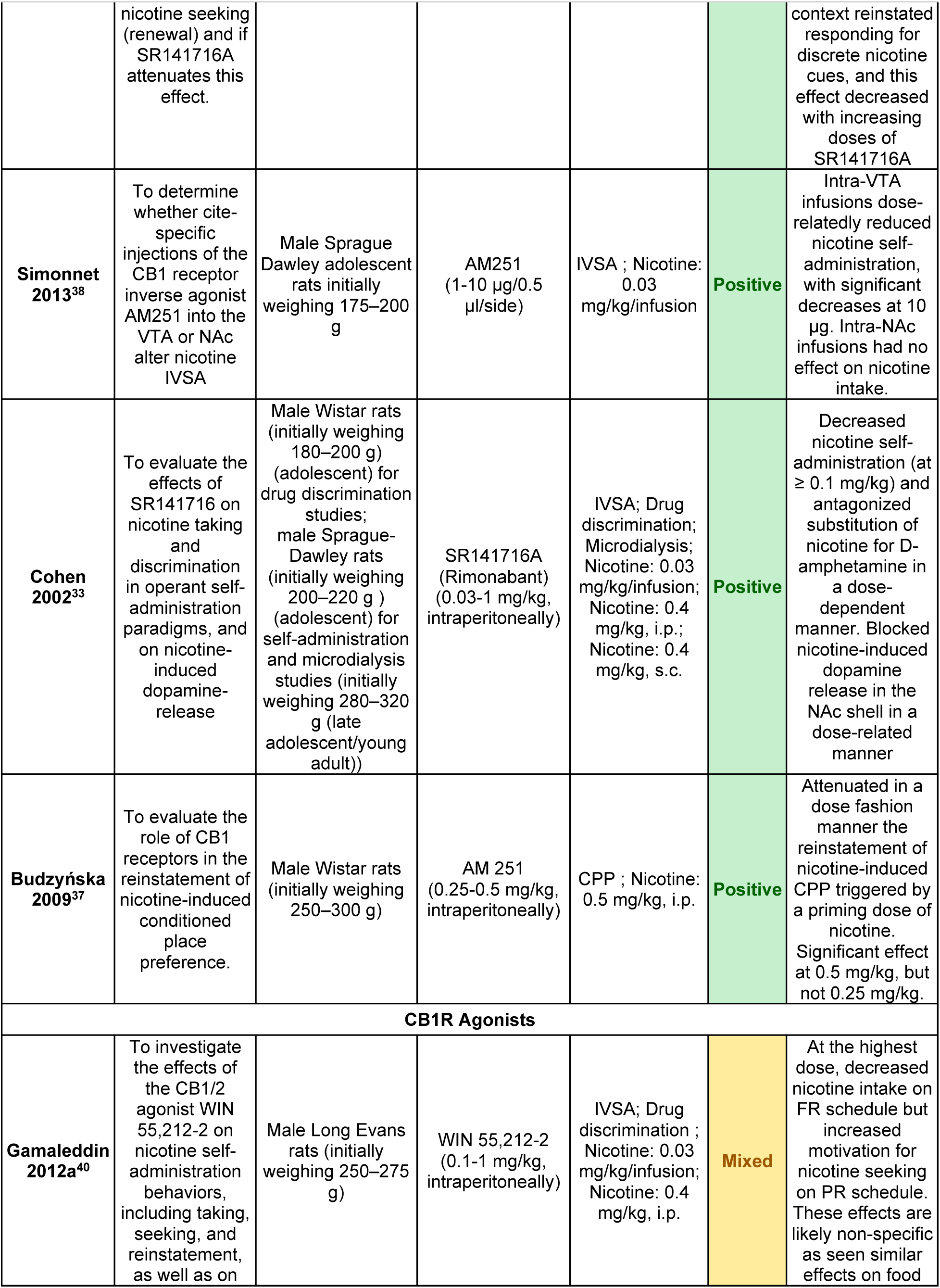
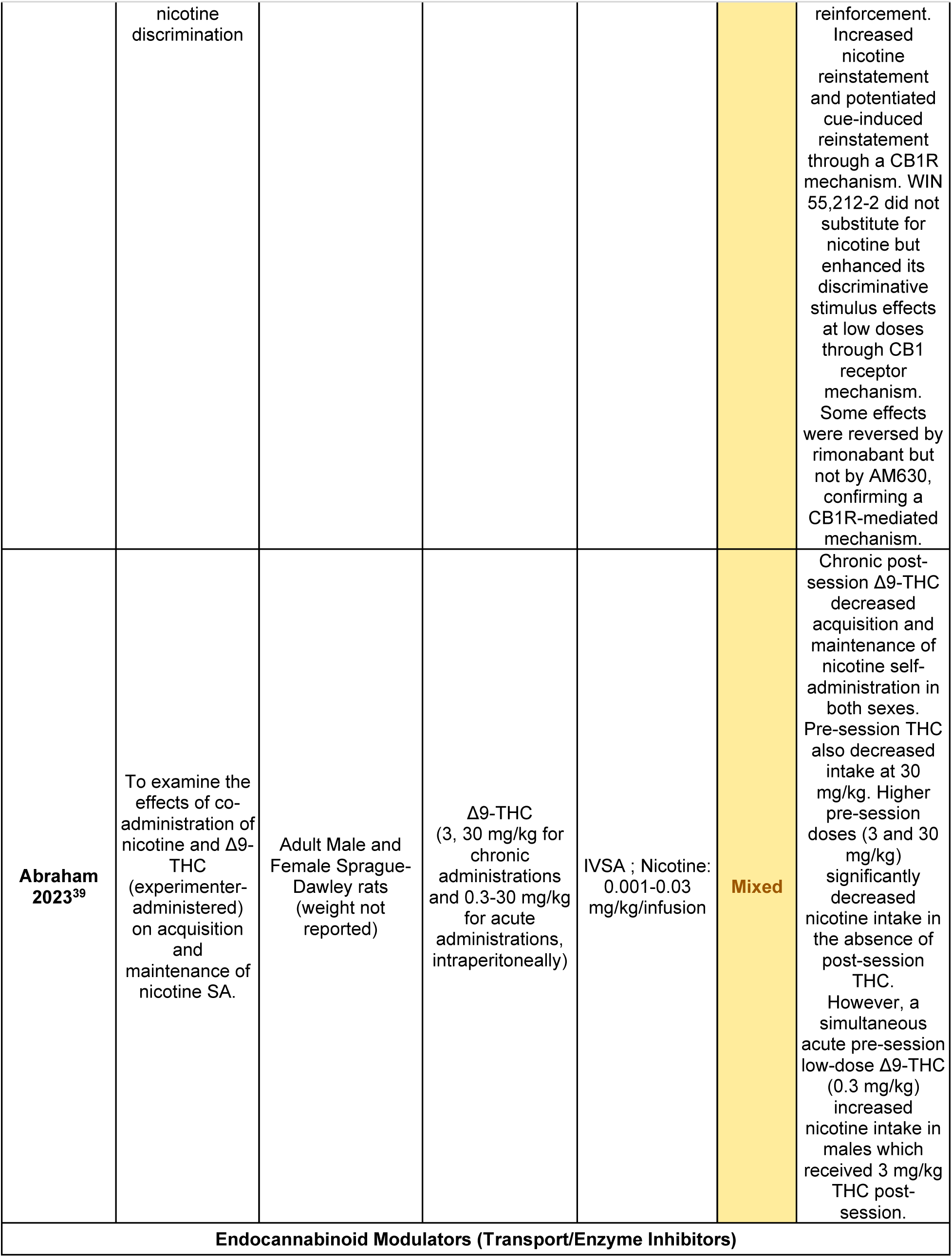
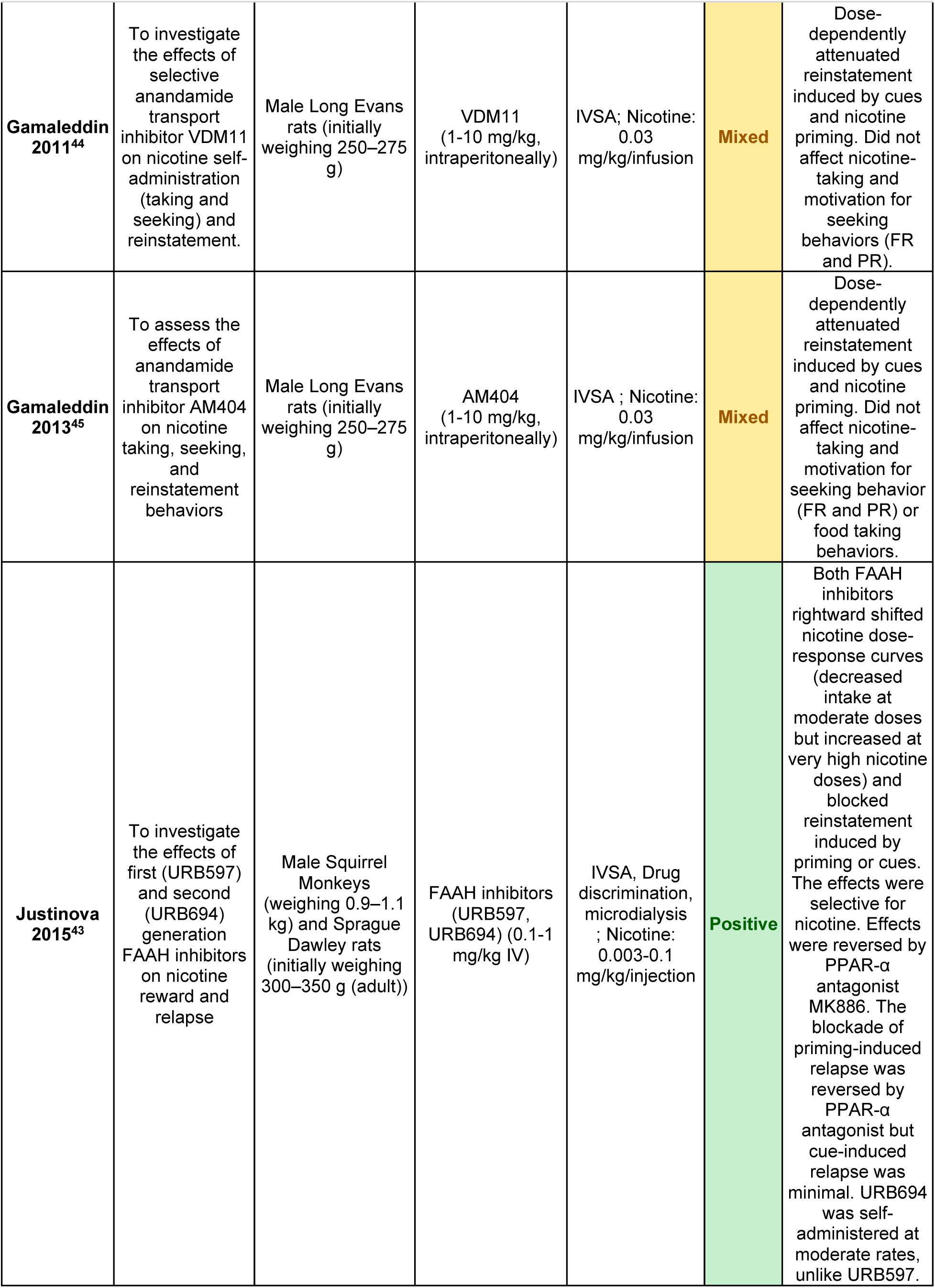
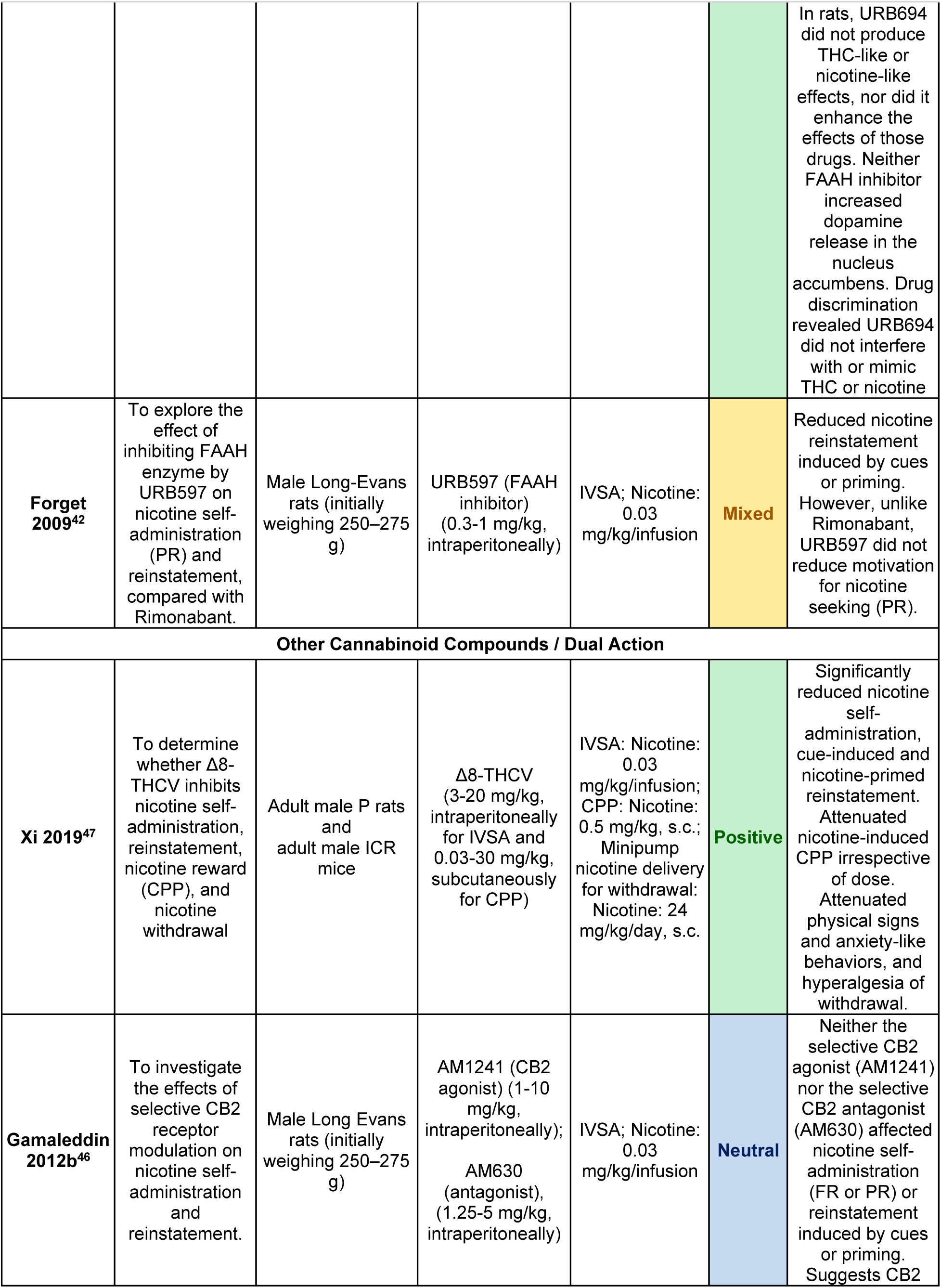
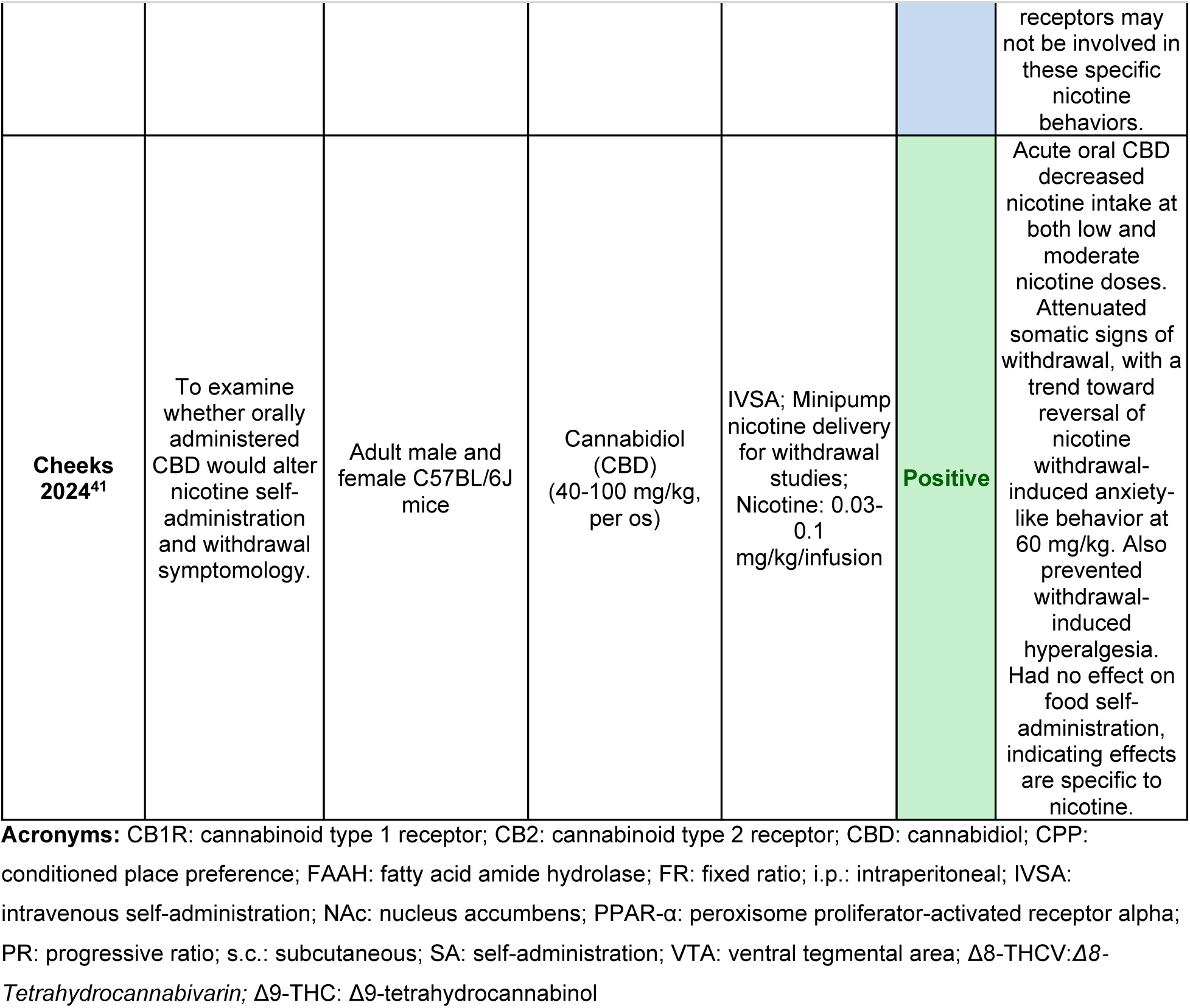
Preclinical studies investigating the effects of CB1R antagonists, inverse agonists, CB1R agonists, endocannabinoid modulators, and other cannabinoid compounds on nicotine use behaviors.

#### 3.2.1 Cannabinoid 1 Receptor Antagonists/Inverse Agonists

Six preclinical studies examined compounds that reduce CB1R signaling, including inverse agonists (AM251, rimonabant/SR141716A), which not only block receptor activation but also suppress constitutive CB1R signaling, and the neutral antagonist SLV330, all of which demonstrated uniformly positive effects^33–38^. The AM251 reduced nicotine self-administration in a dose-dependent manner, with nicotine self-administration showing greater sensitivity to AM251 dose than food self-administration, as indicated by significant reductions at 1 and 3 mg/kg, complete blockade at 10 mg/kg. Food self-administration also trended toward reduction at 1 and 3 mg/kg, though this did not reach significance, with only a partial reduction at the highest dose^34^. In addition to nicotine taking, AM251 also reduced nicotine reinstatement in a dose-dependent manner^34^. Rimonabant decreased nicotine self-administration and additionally blocked nicotine-induced dopamine release in the NAc shell and bed nucleus of the stria terminalis, providing mechanistic support for behavioral observations^33^. Regional specificity studies demonstrated that intra-VTA, but not intra-NAc, AM251 infusions attenuated self-administration, indicating that CB1Rs in the VTA, but not in the NAc, control nicotine self-administration^38^.

The neutral antagonist SLV330, which was developed to improve safety profiles by blocking agonist effects without suppressing baseline receptor activity, reduced both nicotine self-administration and cue-induced reinstatement, showing consistency with the above mentioned ligands^36^. CB1R blockade also consistently attenuated reinstatement, including context-induced renewal^35^ and priming-induced reinstatement of conditioned place preference^37^.

#### 3.2.2 Cannabinoid 1 Receptor Agonists

Two preclinical studies examined CB1R agonists, revealing more complex and context-dependent effects^39,40^. The synthetic CB1/CB2 agonist WIN 55,212-2 produced schedule-dependent effects on nicotine self-administration: decreasing intake under fixed-ratio schedules but significantly increasing breakpoints under progressive-ratio schedules, indicating enhanced motivation for nicotine^40^. However, these effects are likely non-specific, as similar effects were observed on food reinforcement^40^. Although motor impairment was not rigorously assessed (e.g., via rotarod or locomotor activity tests), the parallel reduction in food-reinforced responding at the same doses may suggest that the decrease in nicotine intake under fixed-ratio schedules likely reflects non-specific behavioral suppression, possibly due to motor impairment at higher doses, rather than a selective effect on nicotine reinforcement.

Δ9-THC produced similarly complex results: 3h post-session administration decreased nicotine intake during acquisition and maintenance, shifted dose-response curves downward, and delayed acquisition of stable self-administration^39^. However, these effects were time-dependent and did not generalize to sucrose reinforcement, suggesting a specific nicotine-cannabinoid interaction rather than non-specific behavioral suppression. The effect of pre-session THC on nicotine intake depended heavily on the daily (post-session) THC dose the rats received throughout the study. Higher pre-session doses (3 and 30 mg/kg) significantly decreased nicotine intake when the rats did not receive any post-session administration. When rats in the moderate daily THC group (3 mg/kg post-session) received a low pre-session dose (0.3 mg/kg), nicotine intake increased, particularly in males, an effect not observed in rats that did not receive daily THC. However, the highest pre-session dose (30 mg/kg) still suppressed their nicotine intake. In addition, rats receiving 30 mg/kg during post-session dose showed a general suppression of responding for nicotine regardless of which pre-session THC dose they were given^39^.

#### 3.2.3 Cannabidiol

A single study examined CBD and found that its oral administration dose-dependently decreased intravenous nicotine self-administration in mice at both low (0.03 mg/kg/infusion) and moderate (0.1 mg/kg/infusion) nicotine doses, with significant reductions observed at both 40 and 100 mg/kg CBD without a dose relation^41^. The CBD effects were specific to nicotine reinforcement, with no effect on operant responding for food pellets.

CBD also reduced the severity of some withdrawal symptomology. Following chronic nicotine exposure via osmotic minipump, CBD (30-60 mg/kg) attenuated somatic withdrawal signs and reversed withdrawal-induced hyperalgesia, with a non-significant trend toward reduced anxiety-like behavior at 60 mg/kg in the light-dark box without affecting locomotor activity.

#### 3.2.4 Indirect Cannabinoid System Modulation and Cannabinoid 2 Receptor Ligands

Six studies examined alternative approaches to cannabinoid system modulation, including fatty acid amide hydrolase (FAAH) inhibitors (2 studies)^42,43^, anandamide transport inhibitors (2 studies)^44,45^, selective CB2 receptor ligands (1 study)^46^, and dual-mechanism compounds (1 study)^47^. These investigations reveal mechanism-specific dissociations between effects on nicotine-taking versus nicotine-seeking behavior.

FAAH inhibitors (URB597, URB694) and anandamide transport inhibitors (AM404, VDM11) consistently attenuated cue-induced and nicotine-primed reinstatement without affecting breakpoints (motivation for nicotine seeking) or ongoing self-administration (nicotine taking) in rats^42,44–46^. In squirrel monkeys, FAAH inhibition shifted nicotine dose-response curves rightward and blocked nicotine-primed reinstatement, effects reversed by the PPAR-α antagonist MK886, implicating non-cannabinoid fatty acid ethanolamides rather than anandamide-mediated CB1R activation^43^.

Selective CB2 receptor modulation produced null effects: neither the agonist AM1241 nor antagonist AM630 affected nicotine self-administration or reinstatement^46^.

Δ8-tetrahydrocannabivarin, which combines CB1R antagonist and CB2R agonist properties, demonstrated comprehensive efficacy, attenuating self-administration, cue- and nicotine-primed reinstatement, conditioned place preference acquisition, and all three withdrawal measures (anxiety-like behavior, somatic signs, hyperalgesia) without producing conditioned place preference or aversion and without affecting locomotor activity^47^.

In summary, CB1R antagonism and indirect endocannabinoid tone modulation consistently reduced nicotine reinforcement and relapse across paradigms, while CB1R agonism produced complex, context-dependent effects. CBD reduced both self-administration and withdrawal signs without affecting food reinforcement, and CB2R-selective compounds were ineffective.

### 3.3 Endocannabinoid System Modulation of Nicotine-Related Behaviors: Clinical Evidence

Nineteen human studies examined endocannabinoid system modulation for nicotine addiction, comprising a pooled analysis and individual trials of CB1R inverse agonists (rimonabant, surinabant, taranabant) and six studies of CBD. These investigations employed diverse methodological approaches including randomized controlled trials (parallel-group and crossover), and open-label studies (**Table 2**).

**Table 2.**
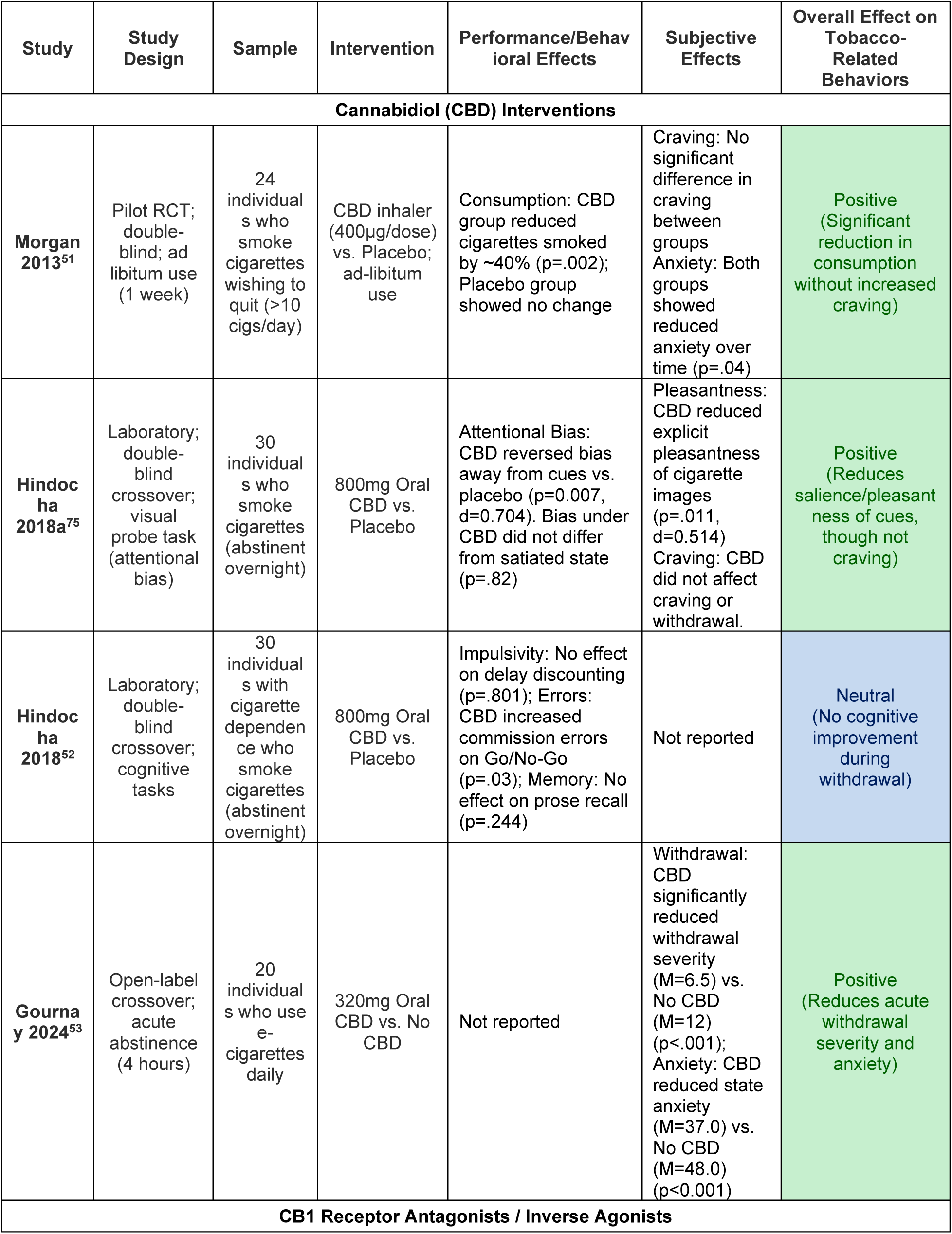
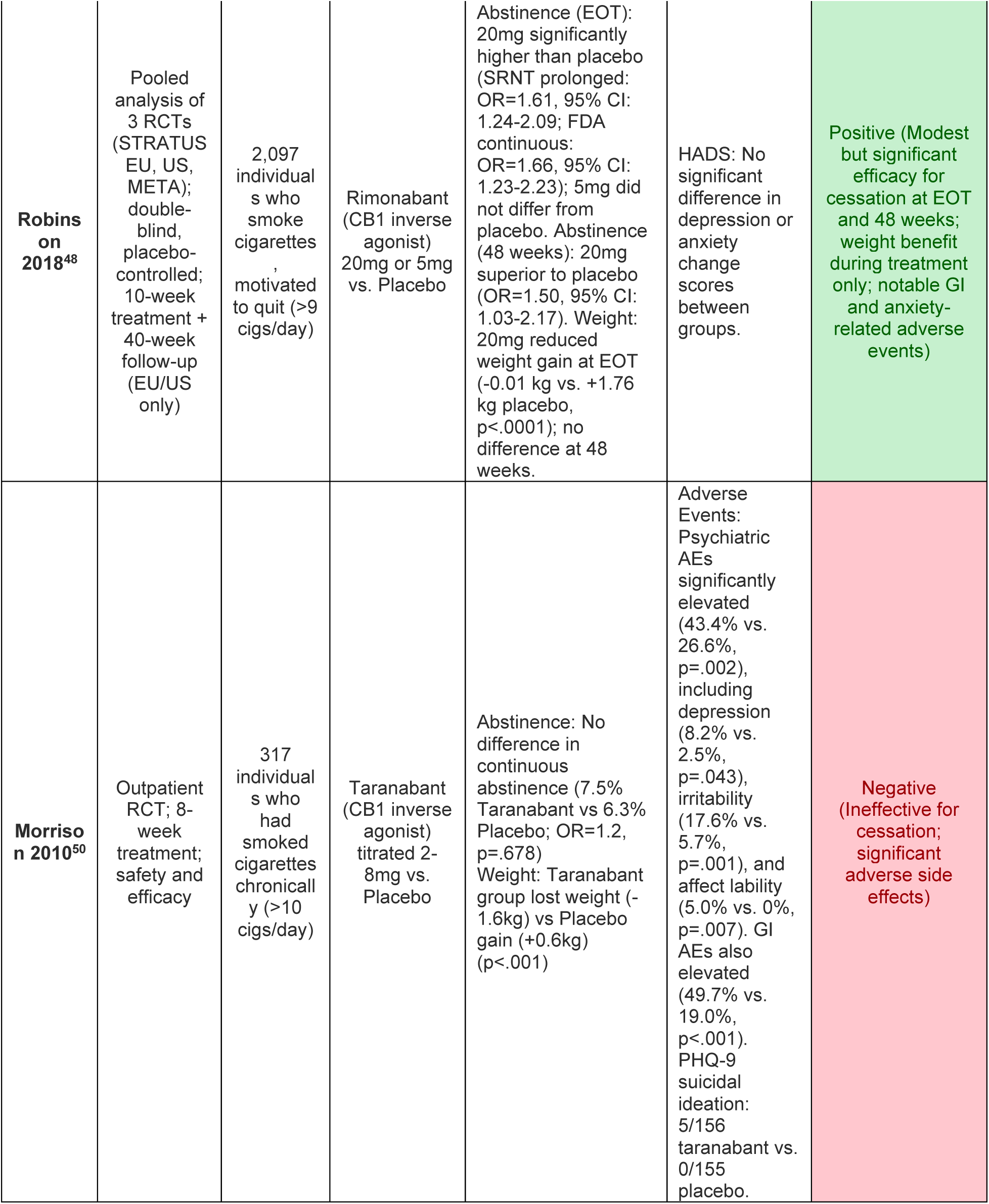
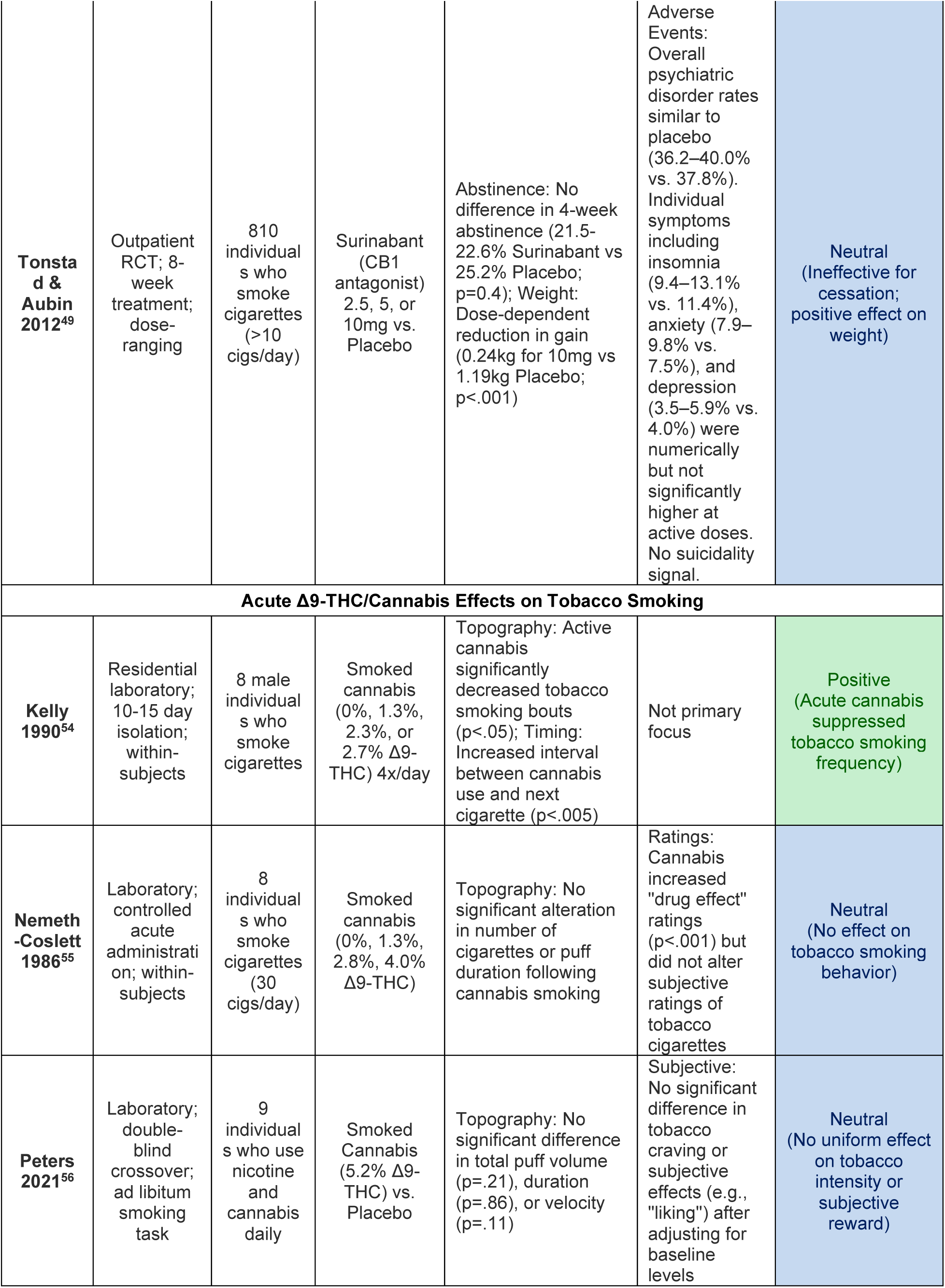
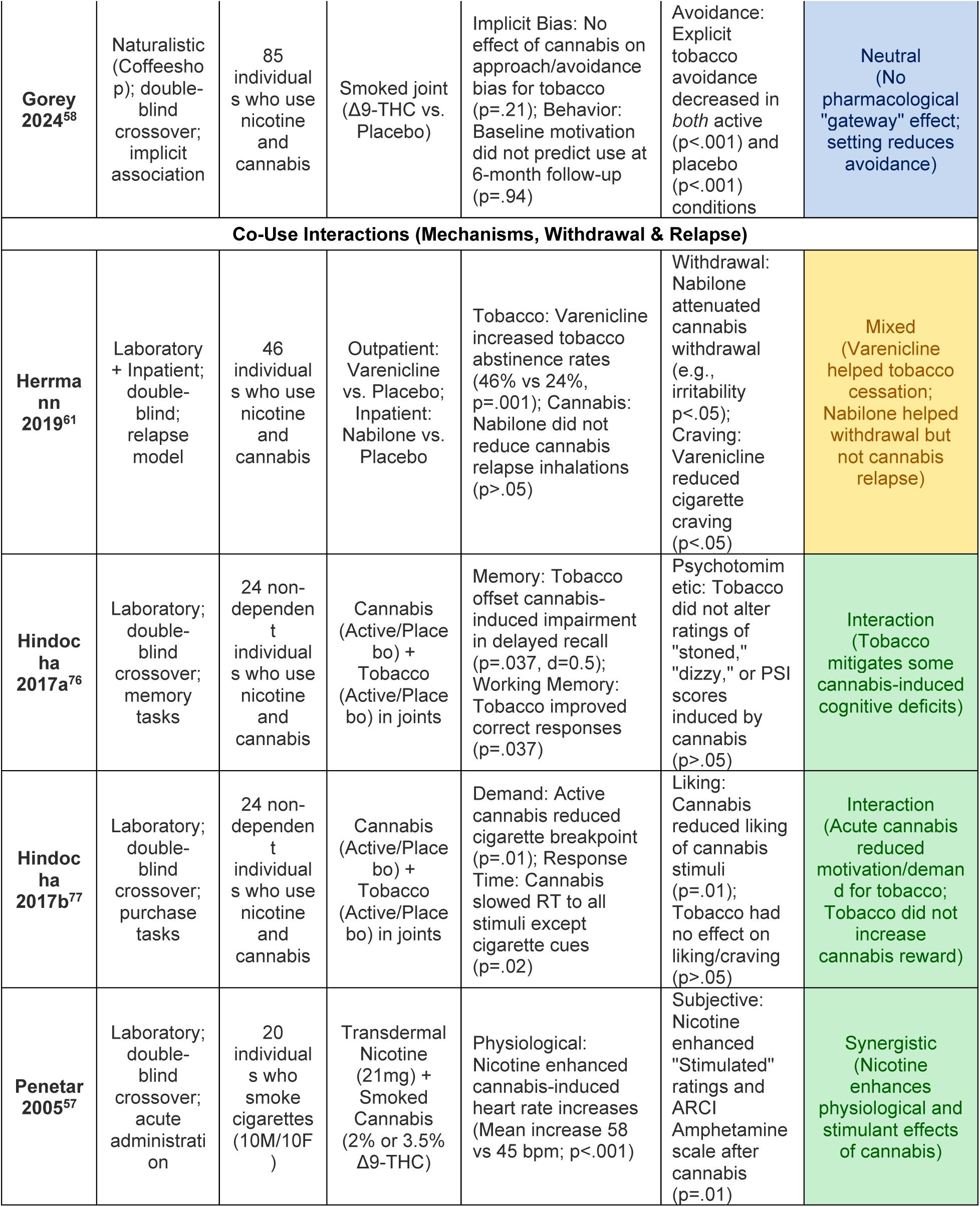
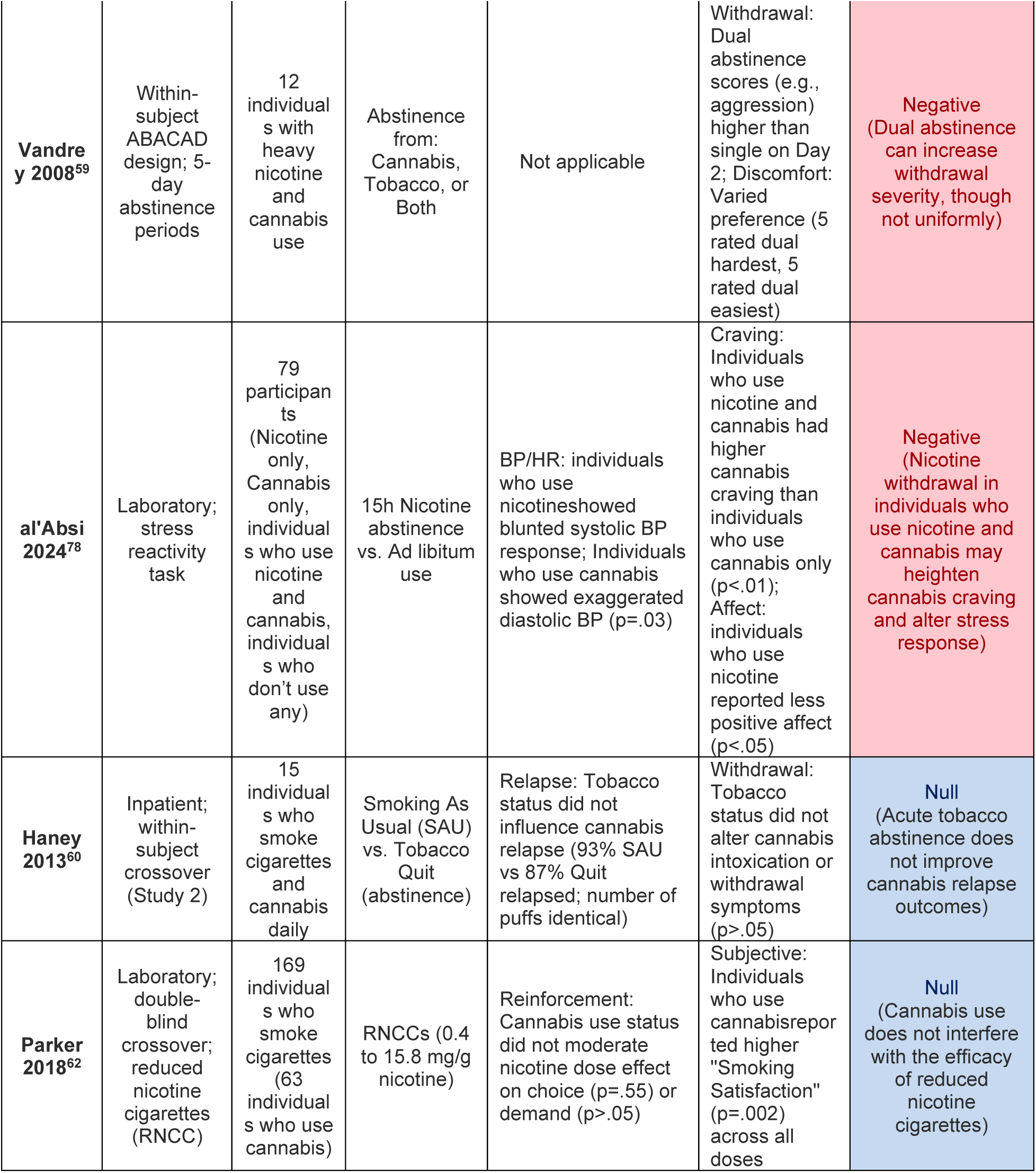
Clinical studies investigating the effects of CB1R inverse agonists, cannabidiol, and acute cannabis-tobacco interactions on tobacco-related outcomes.

#### 3.3.1 Cannabinoid 1 Receptor Inverse Agonists

Three CB1R inverse agonists (rimonabant, surinabant, and taranabant) underwent clinical evaluation for tobacco cessation. A pooled analysis of three rimonabant trials^48^ (STRATUS EU, US, META; N=2,097) found that, at 8-week end-of-treatment, rimonabant 20 mg demonstrated significantly higher prolonged abstinence versus placebo: 21.6% versus 14.7% (OR = 1.61; 95% CI: 1.24, 2.08; p = 0.0004). This advantage persisted at 48-week follow-up: 14.8% versus 10.4% (OR = 1.50; 95% CI: 1.03, 2.17; p = 0.04). The 5 mg dose showed no superiority (OR = 1.05; p = 0.81). The adverse event profile revealed clinically significant concerns: overall events occurred in 69.6% versus 61.0% of placebo recipients, driven by gastrointestinal symptoms and psychiatric manifestations (anxiety: 9.8% vs. 5.7%). Treatment discontinuation rates doubled with rimonabant (11.0% vs. 5.6%; p < 0.001), and post-marketing surveillance ultimately identified serious psychiatric risks including depression and suicidality, leading to market withdrawal in 2008.

A randomized-controlled trial evaluated surinabant (2.5-10 mg/day) in 810 participants over 8 weeks^49^. Four-week continuous abstinence rates showed no dose-response: 22.6%, 22.1%, 21.5% for 2.5, 5, 10 mg versus 25.2% placebo (p trend = 0.40). Surinabant produced dose-dependent weight suppression (0.24-0.75 kg vs. 1.19 kg placebo; p < 0.001). Adverse events reported more frequently with surinabant than placebo included headache (12.9–18.0%), nausea (6.9–14.1%), insomnia (9.4–13.1%), and anxiety (7.9–9.8%), although overall psychiatric disorder rates did not differ meaningfully from placebo (36.2–40.0% vs. 37.8%) and there was no signal for suicidality. Similarly, taranabant (2-8 mg/day titration; N=317) failed to achieve efficacy^50^, with continuous abstinence rates of 7.5% versus 6.3% placebo (OR = 1.2; p = 0.678) despite producing significant weight loss and reduced cigarette consumption among non-abstainers. Taranabant was associated with significantly elevated psychiatric adverse events (43.4% vs. 26.6%; p = 0.002), including depression (8.2% vs. 2.5%; p = 0.043), irritability (17.6% vs. 5.7%; p = 0.001), and affect lability (5.0% vs. 0%; p = 0.007). Five participants in the taranabant group endorsed suicidal ideation on the PHQ-9 versus none receiving placebo, though no suicidal behavior was reported.

#### 3.3.2 Cannabidiol

Four studies evaluated CBD for TUD using diverse methodologies. A double-blind, placebo-controlled crossover trial in 24 daily tobacco smokers (≥10 cigarettes/day) examined CBD inhaler (400 μg/actuation) versus placebo for one week with ad libitum use^51^. The CBD group demonstrated ∼40% reduction in cigarette consumption (p = 0.002), maintained at 2-week follow-up (p = 0.034). Smoking craving assessed by the Tiffany Craving Questionnaire did not differ between groups.

A subsequent crossover study examined single-dose oral CBD 800 mg effects in 30 overnight-abstinent smokers across multiple cognitive and motivational domains^52^. Attentional bias assessment, using a visual probe task, revealed that placebo-treated and recently abstinent tobacco smokers showed marked bias toward cigarette cues at 200 ms exposure, compared to a satiated state (p = 0.001; d = 0.789). CBD administration produced a comparable effect, such that attentional bias did not differ from the satiated state (p = 0.82) and was smaller than that of placebo (p = 0.007). CBD also reduced explicit pleasantness ratings of cigarette images (p = 0.011; d = 0.514), but did not affect self-reported craving or withdrawal symptoms. Cognitive performance assessments revealed null effects on delay discounting (p = 0.801), prose recall (p = 0.244), and n-back tasks (p = 0.472).

Most recently, an open-label crossover study evaluated CBD 320 mg for e-cigarette cessation in 20 individuals who use tobacco daily experiencing anxiety during withdrawal^53^. Following 4-hour supervised abstinence, CBD significantly reduced withdrawal severity (MTWS-R: 6.5 vs. 12.0; *p* < 0.001; *d* = 0.704) and state anxiety (STAI-S: 37.0 vs. 48.0; *p* < 0.001; d=0.514), with large effect sizes. Effects remained significant after controlling for expectancies. CBD was well-tolerated with mild adverse events (somnolence, nausea, headache) in 4 participants.

#### 3.3.3 Human Experimental Evidence on Tobacco-Cannabis Interactions

Beyond targeted pharmacotherapy, a separate body of human experimental research has examined how naturalistic, whole-plant cannabis and tobacco co-use interact, providing mechanistic context for the observational findings in Section 3.1. We organized this literature into three domains: acute behavioral interactions, cognitive and affective mechanisms, and clinical implications for withdrawal and relapse.

##### 3.3.3.1 Acute Behavioral and Subjective Effects

Human experimental research examining whether cannabis use acutely alters tobacco smoking behavior (“chasing”) has produced mixed results. A residential study found that active cannabis smoking significantly decreased the number of daily tobacco smoking bouts but increased the intensity of smoking within bouts (decreased inter-puff intervals)^54^. However, subsequent laboratory studies have generally not supported a strong pharmacological drive for this behavior. Various doses of active cannabis produced no reliable changes in the number of tobacco cigarettes smoked or smoking topography compared to placebo^55^. Similarly, a study on daily people who co-use both substances found that smoking active cannabis did not significantly alter tobacco cigarette craving or puff topography (e.g., puff volume, velocity) compared to placebo, suggesting that the “chasing” phenomenon may be driven more by learning or environmental cues than direct pharmacological interaction^56^.

Regarding subjective and physiological effects, nicotine appears to enhance certain responses to cannabis. Pretreatment with a 21-mg transdermal nicotine patch significantly enhanced cannabis-induced increases in heart rate and subjective ratings of stimulation, particularly in male participants^57^. Conversely, regarding the potential for acute cannabis intoxication to act as a “gateway” via motivational changes, a study found that while cannabis intoxication reduced motivation for cannabis (satiation), it did not increase implicit or explicit motivation for tobacco^58^.

##### 3.3.3.2 Clinical Tobacco Use Disorder Outcomes

A within-subject study compared withdrawal from cannabis, tobacco, and both substances simultaneously. Simultaneous withdrawal produced more severe anger and aggression than withdrawal from either substance alone, while the overall magnitude of cannabis-only withdrawal was comparable to tobacco-only withdrawal^59^.

Two human laboratory studies examined bidirectional relapse dynamics between cannabis and tobacco. People who smoked cannabis daily and also used tobacco cigarettes were more likely to relapse to cannabis use during a laboratory abstinence phase than those who did not smoke cigarettes ^60^. Acute tobacco abstinence did not alter cannabis relapse rates compared to smoking as usual ^60^. Consistent with this asymmetry, a combined laboratory/inpatient study of 46 individuals who co-use nicotine and cannabis found that varenicline increased tobacco abstinence rates (46% vs. 24%; p = .001), while nabilone, a synthetic cannabinoid agonist, attenuated cannabis withdrawal symptoms (e.g., irritability; p < .05) but did not reduce cannabis relapse ^61^.

A secondary analysis of a multi-site, double-blind, laboratory study found that cannabis use status did not moderate the effects of reduced nicotine content cigarettes (RNCC); people who use cannabis and those who do not responded similarly to RNCCs in terms of reduced reinforcement and withdrawal suppression, suggesting that policy-level interventions reducing nicotine content may be equitable across cannabis-using and non-using populations^62^.

In summary, acute co-administration does not consistently show to potentiate nicotine reinforcement, but simultaneous withdrawal from both substances appears to amplify negative affect, and tobacco smoking status predicts cannabis relapse. Reduced nicotine content cigarettes appear equally effective regardless of cannabis use status.

## 4. Discussion

In this translational systematic review and meta-analysis, we synthesized and integrated observational, preclinical, and human experimental evidence to clarify the relationship between cannabinoid signaling and nicotine dependence. Four principal findings emerged. First, observational studies indicate that cannabis co-use is associated with a 35% reduction in the odds of tobacco cessation. Second, preclinical data suggested efficacy signals for certain CB1R inverse agonists; however, clinical translation has thus far been unsuccessful. Although rimonabant demonstrated tobacco cessation efficacy, it was withdrawn due to anxiety, depression, and suicidality, while taranabant and surinabant failed to show cessation benefit, with taranabant additionally linked to depression, irritability, and affective lability. Third, CBD emerges as a promising alternative, given its favorable safety and mechanistic profile. Fourth, human laboratory studies of cannabis–tobacco co-use reveal bidirectional effects on withdrawal and reinforcement, offering a plausible explanation for the association between cannabis co-use and lower tobacco cessation rates.

### 4.1. Implications to Understand Tobacco and Cannabis Co-Use Patterns and Cessation

The 35% reduction in cessation odds among individuals receiving tobacco cessation intervention who co-use cannabis (OR=0.65; 95% CI: 0.55-0.78) represents a clinically meaningful effect, particularly given rising co-use prevalence in the context of expanding cannabis legalization. Notably, the magnitude of this association exceeds that of several established negative prognostic factors in large cessation trials, including anxiety disorder (OR=0.71), psychotic disorder (OR=0.73), and prior failed pharmacotherapy attempts (OR=0.78)^63^, thereby supporting routine assessment of cannabis use status alongside these risk markers in cessation treatment planning.

At the population level, cannabis-tobacco co-use has increased steadily, rising from approximately 4.4% to 6.2% between 2002 and 2019, and has remained elevated through 2023^64,65^. If the association identified here remains stable as co-use prevalence increases, the population-attributable impact on tobacco control may grow correspondingly, particularly given that people who co-use often present with greater dependence severity and more failed quit attempts at baseline^12,13^.

Methodologically, this pooled observational association should be interpreted as robust, but not causal. Observational designs cannot fully adjudicate directionality (e.g., whether cannabis use directly impairs cessation versus marking a higher-risk phenotype), and residual confounding likely contributes. Indeed, people who co-use cannabis and tobacco tend to be younger and male, and present with higher rates of psychiatric comorbidity and polysubstance use^6,11,12^, a clinical profile associated with poorer cessation outcomes^6^. As detailed in **Supplementary Table S2**, while most included studies adjusted for key demographic variables and nicotine dependence severity, few accounted for impulsivity or externalizing psychopathology, and none stratified by cannabis use frequency or quantity. The observed 35% reduction in cessation odds may, therefore, partly reflect a shared vulnerability phenotype rather than a direct pharmacological effect of cannabis on cessation. Still, the consistency of associations across extensively adjusted studies suggests confounding does not fully explain the signal.

Mechanistically, several pathways plausibly link co-use to cessation difficulty, though direct pharmacological effects of Δ9-THC on nicotine reinforcement remain inconclusive (one preclinical study reported enhancement^66^; another found reduction^39^; human laboratory studies have generally not confirmed potentiation). More consistently supported mechanisms include withdrawal amplification, dual abstinence from cannabis and tobacco may intensify negative affect^59^, and nicotine dependence may heighten cannabis withdrawal severity^67^, and conditioned co-administration, where concurrent use of cannabis/tobacco within the same product creates associative cues that trigger relapse to either drug^10^.

Clinically, these findings support routine cannabis use assessment in tobacco treatment settings and treatment plans that anticipate bidirectional craving and withdrawal dynamics.

### 4.2 Emerging Endocannabinoid Targets for Tobacco Cessation

While preclinical evidence robustly supports CB1R antagonism, with all six studies reporting favorable effects with regional specificity and mechanistic confirmation, clinical translation has failed. Despite demonstrating biological activity taranabant and surinabant showed no cessation efficacy. Taranabant was associated with significantly elevated psychiatric adverse events, including depression and irritability, while surinabant’s psychiatric adverse event rates did not differ meaningfully from placebo, though individual symptoms such as insomnia and anxiety were numerically more frequent at higher doses^49,50^. These outcomes may be specific to inverse agonism rather than competitive antagonism, suggesting that CB1R target remains valid but the pharmacological approach requires refinement. For instance, through neutral antagonists. As a result, the field has shifted toward ECS modulation strategies that bypass direct CB1R blockade.

CBD emerges as the most clinically viable treatment approach targeting the ECS for TUD. Preliminary proof-of-concept^51^ is provided by Morgan and colleagues, who observed 40% reduction in cigarette consumption, though this requires replication in adequately powered, rigorously controlled trials. Unlike direct CB1R inverse agonists, CBD engages multiple addiction-relevant pathways^68^: negative allosteric modulation of CB1R may attenuate reward signaling without suppressing constitutive receptor activity^69^, thereby avoiding the depressive liability of inverse agonism; 5-HT1A agonism may underlie the anxiolytic and anti-craving effects observed during acute withdrawal^53,70^; TRPV1 activation contributes to modulation of mesolimbic dopamine transmission^71,72^; and FAAH inhibition enhances endocannabinoid tone, which has independently demonstrated anti-reinstatement efficacy in preclinical nicotine models^42,43,73^. Additionally, CBD inhibits nicotine metabolism via CYP2A6^74^, potentially prolonging nicotine exposure per cigarette and reducing compensatory smoking, a pharmacokinetic mechanism distinct from its receptor-level effects.

Beyond CBD, minor cannabinoids represent additional emerging targets. Δ8-tetrahydrocannabivarin (Δ8-THCV), which combines CB1R antagonist and CB2R agonist properties, reduced nicotine self-administration preclinically without the psychiatric adverse effect profile of CB1R inverse agonists^47^. FAAH inhibitors and anandamide transport inhibitors similarly attenuated reinstatement-like behaviors in preclinical models^42,44^, pointing toward AEA enhancement as a viable mechanistic strategy. Collectively, these emerging targets share a common principle: indirect or partial modulation of the ECS may achieve therapeutic benefit without the liabilities of direct, full CB1R blockade.

Methodologically, advancing CBD (and related ECS tone-modulating approaches) will require careful attention to product characterization (composition/contaminants), dose selection anchored in exposure-response, and endpoint selection that aligns with mechanism (e.g., cue reactivity, reinforcement, withdrawal coupling), ideally alongside biochemical verification of tobacco and cannabis use.

### 4.3 Limitations

Several limitations warrant consideration. First, the meta-analysis revealed high heterogeneity (I² = 88.1%) across studies, likely reflecting differences in cannabis use definitions, cessation outcome measures, follow-up durations, and abstinence verification methods. Furthermore, this heterogeneity captures the clinical diversity of the included studies (**Supplementary Table S2**), which used different interventions (ranging from pharmacotherapy to digital health tools), targeted distinct populations (e.g., adolescents, adults with diabetes), and included varied nicotine products (combustible cigarettes vs. e-cigarettes). Second, although formal publication bias tests (Egger’s p = 0.071; Begg’s p = 0.948) were statistically non-significant, selective reporting of positive findings cannot be excluded. Third, observational designs preclude causal inference, as unmeasured confounders, including psychiatric comorbidity, polysubstance use, and nicotine dependence severity, may partially account for the observed association. Fourth, most included studies did not biochemically verify cannabis use or smoking abstinence, introducing potential misclassification bias. Fifth, nearly all observational studies assessed cannabis use as a binary variable (past 30-day any use vs. none), with route of administration unreported in most cases. This precludes dose-response analysis and limits the distinction of pharmacological effects of cannabis use from shared vulnerability.

Additional limitations should be considered when interpreting the preclinical literature. Female subjects were often not included, and age was not reported, limiting conclusions regarding sex differences and developmental stage (adolescent versus adult). In addition, some studies lacked appropriate controls, such as alternative reinforcers (e.g., food) or positive control compounds, making it difficult to determine whether effects were specific to nicotine.

Finally, many studies did not directly assess potential motor impairments at higher doses and instead relied on speculation to explain non-specific behavioral effects.

### 4.4 Future Directions

A central translational paradox emerges from this review: naturalistic cannabis co-use is associated with 35% lower cessation odds, yet targeted ECS modulation, particularly CBD, shows therapeutic promise. Three variables likely account for this divergence: (1) ligand composition, as naturalistic cannabis delivers Δ9-THC alongside other phytocannabinoids with widely varying potency and Δ9-THC-to-CBD ratios^66^, whereas CBD engages multiple receptor systems without Δ9-THC’s bidirectional effects on nicotine reinforcement^39,66^; (2) pharmacokinetics, as smoked cannabis produces rapid Δ9-THC peaks that may promote conditioned co-administration with tobacco^10^, whereas oral formulations yield slower, sustained exposure better suited to therapeutic contexts^51,53^; and (3) neuroadaptation state, as simultaneous withdrawal amplifies negative affect beyond either substance alone^59^, compounding cessation difficulty among people who co-use who already present with greater dependence severity^12,13^. These hypothesized sources of divergence inform three synergistic research priorities.

First, elucidating the mechanisms by which co-use impairs cessation across addiction stages. The observational association (OR = 0.65) likely reflects contributions from reinforcement, withdrawal, and relapse, but the relative importance of each remains unclear. Human laboratory studies should isolate whether co-use primarily amplifies withdrawal severity, enhances cue-conditioned relapse, or alters reinforcement processing, ideally using paradigms that distinguish these stages within the same individuals. Prospective observational studies with granular cannabis exposure characterization (frequency, quantity, product type, Δ9-THC-to-CBD ratio, route of administration) are needed to move beyond the binary measures that currently preclude dose-response analysis. This is a critical gap because concentration of the association among people who use cannabis heavily would help distinguish cannabis-specific mechanisms from shared vulnerability.

Second, systematic evaluation of emerging ECS targets. Adequately powered randomized controlled trials of CBD for tobacco cessation, examining dose-response relationships, optimal formulations (inhaled vs. oral), and treatment duration, represent a high priority. Δ8-THCV^47^ and FAAH inhibitors^42,43^ also merit continued development given consistent preclinical efficacy and mechanistic distinctiveness from failed inverse agonists. The conflicting preclinical findings on Δ9-THC and nicotine reinforcement need resolution through parametric designs systematically varying dose, timing, and chronicity of prior cannabinoid exposure.

Additionally, prospective studies should examine whether cannabis co-use moderates response to existing cessation pharmacotherapies (varenicline, NRT); if people who co-use respond differently, tailored treatment algorithms may be warranted.

Third, developing clinical guidance for managing tobacco cessation in people with co-use. This represents the most immediate translational gap: the current evidence establishes that co-use predicts poorer outcomes but provides no management guidance. Key unanswered questions include whether co-use should prompt concurrent treatment of both substances versus sequential approaches; whether routine screening for cannabis use, with attention to frequency, route, and product type, should be incorporated into standard cessation assessments alongside established risk markers such as psychiatric comorbidity and prior failed attempts^63^; and how clinicians should counsel people who co-use regarding the distinction between Δ9-THC-dominant products and ECS-targeted pharmacotherapies under clinical investigation. Integrated behavioral interventions addressing bidirectional withdrawal dynamics and conditioned co-administration may also warrant development.

## 5. Conclusions

In this translational systematic review and meta-analysis, we have highlighted the endocannabinoid-nicotinic interface as both a clinical challenge and therapeutic frontier.

We provide evidence that cannabis co-use is associated with 35% lower odds of tobacco cessation success. The mechanistic basis of this association remains incompletely understood and may partly reflect confounding by shared vulnerability factors, rather than direct pharmacological effects. These findings, nonetheless, underscore the need for routine co-use assessment in cessation settings. Simultaneously, convergent preclinical studies show that targeted pharmacological modulation of the ECS shows mechanistic promise, particularly through CBD and minor cannabinoids (e.g., Δ8-THCV) that avoid the rapid reinforcement dynamics of smoked, Δ9-THC-dominant products, and the psychiatric adverse effects of CB1R inverse agonists. Finally, building on the evidence synthesized and appraised, we articulate how distinction between naturalistic cannabinoid exposure and targeted pharmacotherapy, determined by ligand composition, pharmacokinetics, and prior neuroadaptations, has direct implications for both clinical practice and therapeutic development.

## Author Contributions (CRediT)

GPAC: Conceptualization, Methodology, Formal analysis, Data curation, Investigation, Writing – Original Draft, Visualization. OG: Data curation, Investigation, Writing – Review & Editing. LRdR: Data curation, Investigation. MACM: Data curation. MCF: Methodology, Resources, Data curation, Writing – Review & Editing. DB: Supervision, Writing – Review & Editing. MS: Conceptualization, Supervision, Writing – Review & Editing. JPDA: Conceptualization, Methodology, Supervision, Project administration, Writing – Review & Editing. All authors approved the final version of the manuscript and agree to be accountable for all aspects of the work.

## Data Statement

All data generated or analyzed during this study are included in this published article and its supplementary information files. The raw data used for meta-analyses are available from the corresponding author upon reasonable request.

## Declaration of generative AI and AI-assisted technologies in the manuscript preparation process

Prior to manuscript submission, the authors used Claude (Anthropic) to assist in identifying potential typographical errors and internal inconsistencies. The authors reviewed and edited all content as needed and take full responsibility for the accuracy and integrity of the final manuscript.

## Acknowledgements

None.

## References

1. Spatial, temporal, and demographic patterns in prevalence of smoking tobacco use and attributable disease burden in 204 countries and territories, 1990-2019: a systematic analysis from the Global Burden of Disease Study 2019. Lancet. Jun 19 2021;397(10292):2337–2360. doi:10.1016/s0140-6736(21)01169-7

2. Institute for Health M, Evaluation. Global Burden of Disease 2023. [online application]. 2025

3. Cahill K, Stevens S, Perera R, Lancaster T. Pharmacological interventions for smoking cessation: an overview and network meta-analysis. Cochrane Database Syst Rev. May 31 2013;2013(5):Cd009329. doi:10.1002/14651858.CD009329.pub2

4. Hughes JR, Keely J, Naud S. Shape of the relapse curve and long-term abstinence among untreated smokers. Addiction. Jan 2004;99(1):29–38. doi:10.1111/j.1360-0443.2004.00540.x

5. Evins AE, Benowitz NL, West R, et al. Neuropsychiatric Safety and Efficacy of Varenicline, Bupropion, and Nicotine Patch in Smokers With Psychotic, Anxiety, and Mood Disorders in the EAGLES Trial. J Clin Psychopharmacol. Mar/Apr 2019;39(2):108–116. doi:10.1097/jcp.0000000000001015

6. Peters EN, Budney AJ, Carroll KM. Clinical correlates of co-occurring cannabis and tobacco use: a systematic review. Addiction. Aug 2012;107(8):1404–17. doi:10.1111/j.1360-0443.2012.03843.x

7. Costa GPA, Asnes S, De Aquino JP. Tobacco-cannabis co-use in adults ≥50: Trends, medical marijuana laws, and cessation implications-Letter to the Editor. Am J Addict. Nov 2025;34(6):682–683. doi:10.1111/ajad.70088

8. Reboussin BA, Wagoner KG, Ross JC, Suerken CK, Sutfin EL. Tobacco and marijuana co-use in a cohort of young adults: Patterns, correlates and reasons for co-use. Drug Alcohol Depend. Oct 1 2021;227:109000. doi:10.1016/j.drugalcdep.2021.109000

9. Akbar SA, Tomko RL, Salazar CA, Squeglia LM, McClure EA. Tobacco and cannabis co-use and interrelatedness among adults. Addict Behav. Mar 2019;90:354–361. doi:10.1016/j.addbeh.2018.11.036

10. Ream GL, Benoit E, Johnson BD, Dunlap E. Smoking tobacco along with marijuana increases symptoms of cannabis dependence. Drug Alcohol Depend. Jun 1 2008;95(3):199–208. doi:10.1016/j.drugalcdep.2008.01.011

11. Nguyen N, Bold KW, McClure EA. Urgent need for treatment addressing co-use of tobacco and cannabis: An updated review and considerations for future interventions. Addict Behav. Nov 2024;158:108118. doi:10.1016/j.addbeh.2024.108118

12. Agrawal A, Budney AJ, Lynskey MT. The co-occurring use and misuse of cannabis and tobacco: a review. Addiction. Jul 2012;107(7):1221–33. doi:10.1111/j.1360-0443.2012.03837.x

13. Tullis LM, Dupont R, Frost-Pineda K, Gold MS. Marijuana and tobacco: a major connection? J Addict Dis. 2003;22(3):51–62. doi:10.1300/J069v22n03_05

14. Pacher P, Bátkai S, Kunos G. The endocannabinoid system as an emerging target of pharmacotherapy. Pharmacol Rev. Sep 2006;58(3):389–462. doi:10.1124/pr.58.3.2

15. Lu HC, Mackie K. An Introduction to the Endogenous Cannabinoid System. Biol Psychiatry. Apr 1 2016;79(7):516–25. doi:10.1016/j.biopsych.2015.07.028

16. Marsicano G, Lutz B. Expression of the cannabinoid receptor CB1 in distinct neuronal subpopulations in the adult mouse forebrain. Eur J Neurosci. Dec 1999;11(12):4213–25. doi:10.1046/j.1460-9568.1999.00847.x

17. Herkenham M, Lynn AB, Little MD, et al. Cannabinoid receptor localization in brain. Proc Natl Acad Sci U S A. Mar 1990;87(5):1932–6. doi:10.1073/pnas.87.5.1932

18. Parsons LH, Hurd YL. Endocannabinoid signalling in reward and addiction. Nat Rev Neurosci. Oct 2015;16(10):579–94. doi:10.1038/nrn4004

19. Dani JA. Neuronal Nicotinic Acetylcholine Receptor Structure and Function and Response to Nicotine. Int Rev Neurobiol. 2015;124:3–19. doi:10.1016/bs.irn.2015.07.001

20. Changeux JP. Nicotine addiction and nicotinic receptors: lessons from genetically modified mice. Nat Rev Neurosci. Jun 2010;11(6):389–401. doi:10.1038/nrn2849

21. Mansvelder HD, Keath JR, McGehee DS. Synaptic mechanisms underlie nicotine-induced excitability of brain reward areas. Neuron. Mar 14 2002;33(6):905–19. doi:10.1016/s0896-6273(02)00625-6

22. Picciotto MR, Zoli M, Rimondini R, et al. Acetylcholine receptors containing the beta2 subunit are involved in the reinforcing properties of nicotine. Nature. Jan 8 1998;391(6663):173–7. doi:10.1038/34413

23. Buczynski MW, Polis IY, Parsons LH. The volitional nature of nicotine exposure alters anandamide and oleoylethanolamide levels in the ventral tegmental area. Neuropsychopharmacology. Mar 2013;38(4):574–84. doi:10.1038/npp.2012.210

24. Saravia R, Ten-Blanco M, Pereda-Pérez I, Berrendero F. New Insights in the Involvement of the Endocannabinoid System and Natural Cannabinoids in Nicotine Dependence. Int J Mol Sci. Dec 10 2021;22(24)doi:10.3390/ijms222413316

25. Patel JC, Sherpa AD, Melani R, et al. GABA co-released from striatal dopamine axons dampens phasic dopamine release through autoregulatory GABA(A) receptors. Cell Rep. Mar 26 2024;43(3):113834. doi:10.1016/j.celrep.2024.113834

26. Vallés AS, Barrantes FJ. Interactions between the Nicotinic and Endocannabinoid Receptors at the Plasma Membrane. Membranes (Basel). Aug 22 2022;12(8)doi:10.3390/membranes12080812

27. González S, Cascio MG, Fernández-Ruiz J, Fezza F, Di Marzo V, Ramos JA. Changes in endocannabinoid contents in the brain of rats chronically exposed to nicotine, ethanol or cocaine. Brain Res. Nov 1 2002;954(1):73–81. doi:10.1016/s0006-8993(02)03344-9

28. Rabin RA, Farrugia J, Garani R, Mizrahi R, Rusjan P. A preliminary investigation of tobacco co-use on endocannabinoid activity in people with cannabis use. Drug Alcohol Depend Rep. Sep 2025;16:100369. doi:10.1016/j.dadr.2025.100369

29. Scherma M, Masia P, Satta V, Fratta W, Fadda P, Tanda G. Brain activity of anandamide: a rewarding bliss? Acta Pharmacol Sin. Mar 2019;40(3):309–323. doi:10.1038/s41401-018-0075-x

30. Le Foll B. Chapter 11 - Role of anandamide in nicotine addiction. In: Foll BL, ed. Anandamide in Health and Disease. Academic Press; 2025:369–385.

31. Hirvonen J, Zanotti-Fregonara P, Gorelick DA, et al. Decreased Cannabinoid CB(1) Receptors in Male Tobacco Smokers Examined With Positron Emission Tomography. Biol Psychiatry. Nov 15 2018;84(10):715–721. doi:10.1016/j.biopsych.2018.07.009

32. Page MJ, McKenzie JE, Bossuyt PM, et al. The PRISMA 2020 statement: an updated guideline for reporting systematic reviews. BMJ. 2021;372:n71. doi:10.1136/bmj.n71

33. Cohen C, Perrault G, Voltz C, Steinberg R, Soubrié P. SR141716, a central cannabinoid (CB(1)) receptor antagonist, blocks the motivational and dopamine-releasing effects of nicotine in rats. Behav Pharmacol. Sep 2002;13(5-6):451–63. doi:10.1097/00008877-200209000-00018

34. Shoaib M. The cannabinoid antagonist AM251 attenuates nicotine self-administration and nicotine-seeking behaviour in rats. Neuropharmacology. Feb 2008;54(2):438–44. doi:10.1016/j.neuropharm.2007.10.011

35. Diergaarde L, de Vries W, Raasø H, Schoffelmeer AN, De Vries TJ. Contextual renewal of nicotine seeking in rats and its suppression by the cannabinoid-1 receptor antagonist Rimonabant (SR141716A). Neuropharmacology. Oct 2008;55(5):712–6. doi:10.1016/j.neuropharm.2008.06.003

36. de Bruin NM, Lange JH, Kruse CG, et al. SLV330, a cannabinoid CB(1) receptor antagonist, attenuates ethanol and nicotine seeking and improves inhibitory response control in rats. Behav Brain Res. Mar 1 2011;217(2):408–15. doi:10.1016/j.bbr.2010.11.013

37. Budzyńska B, Kruk M, Biała G. Effects of the cannabinoid CB1 receptor antagonist AM 251 on the reinstatement of nicotine-conditioned place preference by drug priming in rats. Pharmacol Rep. Mar–Apr 2009;61(2):304–10. doi:10.1016/s1734-1140(09)70036-2

38. Simonnet A, Cador M, Caille S. Nicotine reinforcement is reduced by cannabinoid CB1 receptor blockade in the ventral tegmental area. Addict Biol. Nov 2013;18(6):930–6. doi:10.1111/j.1369-1600.2012.00476.x

39. Abraham AD, Wiley JL, Marusich JA. Experimenter administered Δ(9)-THC decreases nicotine self-administration in a rat model. Pharmacol Biochem Behav. Oct 2023;231:173632. doi:10.1016/j.pbb.2023.173632

40. Gamaleddin I, Wertheim C, Zhu AZ, et al. Cannabinoid receptor stimulation increases motivation for nicotine and nicotine seeking. Addict Biol. Jan 2012;17(1):47–61. doi:10.1111/j.1369-1600.2011.00314.x

41. Cheeks SN, Buzzi B, Valdez A, Mogul AS, Damaj MI, Fowler CD. Cannabidiol as a potential cessation therapeutic: Effects on intravenous nicotine self-administration and withdrawal symptoms in mice. Neuropharmacology. Mar 15 2024;246:109833. doi:10.1016/j.neuropharm.2023.109833

42. Forget B, Coen KM, Le Foll B. Inhibition of fatty acid amide hydrolase reduces reinstatement of nicotine seeking but not break point for nicotine self-administration--comparison with CB(1) receptor blockade. Psychopharmacology (Berl). Sep 2009;205(4):613–24. doi:10.1007/s00213-009-1569-5

43. Justinova Z, Panlilio LV, Moreno-Sanz G, et al. Effects of Fatty Acid Amide Hydrolase (FAAH) Inhibitors in Non-Human Primate Models of Nicotine Reward and Relapse. Neuropsychopharmacology. Aug 2015;40(9):2185–97. doi:10.1038/npp.2015.62

44. Gamaleddin I, Guranda M, Goldberg SR, Le Foll B. The selective anandamide transport inhibitor VDM11 attenuates reinstatement of nicotine seeking behaviour, but does not affect nicotine intake. Br J Pharmacol. Nov 2011;164(6):1652–60. doi:10.1111/j.1476-5381.2011.01440.x

45. Gamaleddin I, Guranda M, Scherma M, et al. AM404 attenuates reinstatement of nicotine seeking induced by nicotine-associated cues and nicotine priming but does not affect nicotine- and food-taking. J Psychopharmacol. Jun 2013;27(6):564–71. doi:10.1177/0269881113477710

46. Gamaleddin I, Zvonok A, Makriyannis A, Goldberg SR, Le Foll B. Effects of a selective cannabinoid CB2 agonist and antagonist on intravenous nicotine self administration and reinstatement of nicotine seeking. PLoS One. 2012;7(1):e29900. doi:10.1371/journal.pone.0029900

47. Xi ZX, Muldoon P, Wang XF, et al. Δ(8) -Tetrahydrocannabivarin has potent anti-nicotine effects in several rodent models of nicotine dependence. Br J Pharmacol. Dec 2019;176(24):4773–4784. doi:10.1111/bph.14844

48. Robinson JD, Cinciripini PM, Karam-Hage M, et al. Pooled analysis of three randomized, double-blind, placebo controlled trials with rimonabant for smoking cessation. Addict Biol. Jan 2018;23(1):291–303. doi:10.1111/adb.12508

49. Tonstad S, Aubin HJ. Efficacy of a dose range of surinabant, a cannabinoid receptor blocker, for smoking cessation: a randomized controlled clinical trial. J Psychopharmacol. Jul 2012;26(7):1003–9. doi:10.1177/0269881111431623

50. Morrison MF, Ceesay P, Gantz I, Kaufman KD, Lines CR. Randomized, controlled, double-blind trial of taranabant for smoking cessation. Psychopharmacology (Berl). Apr 2010;209(3):245–53. doi:10.1007/s00213-010-1790-2

51. Morgan CJ, Das RK, Joye A, Curran HV, Kamboj SK. Cannabidiol reduces cigarette consumption in tobacco smokers: preliminary findings. Addict Behav. Sep 2013;38(9):2433–6. doi:10.1016/j.addbeh.2013.03.011

52. Hindocha C, Freeman TP, Grabski M, et al. The effects of cannabidiol on impulsivity and memory during abstinence in cigarette dependent smokers. Sci Rep. May 15 2018;8(1):7568. doi:10.1038/s41598-018-25846-2

53. Gournay LR, Petry J, Bilsky S, et al. Cannabidiol Reduces Nicotine Withdrawal Severity and State Anxiety During an Acute E-cigarette Abstinence Period: A Novel, Open-Label Study. Cannabis Cannabinoid Res. Aug 2024;9(4):996–1005. doi:10.1089/can.2022.0317

54. Kelly TH, Foltin RW, Rose AJ, Fischman MW, Brady JV. Smoked marijuana effects on tobacco cigarette smoking behavior. J Pharmacol Exp Ther. Mar 1990;252(3):934–44.

55. Nemeth-Coslett R, Henningfield JE, O’Keeffe MK, Griffiths RR. Effects of marijuana smoking on subjective ratings and tobacco smoking. Pharmacol Biochem Behav. Sep 1986;25(3):659–65. doi:10.1016/0091-3057(86)90156-5

56. Peters EN, Herrmann ES, Smith C, et al. Impact of smoked cannabis on tobacco cigarette smoking intensity and subjective effects: A placebo-controlled, double-blind, within-subjects human laboratory study. Exp Clin Psychopharmacol. Aug 2021;29(4):345–354. doi:10.1037/pha0000391

57. Penetar DM, Kouri EM, Gross MM, et al. Transdermal nicotine alters some of marihuana’s effects in male and female volunteers. Drug Alcohol Depend. Aug 1 2005;79(2):211–23. doi:10.1016/j.drugalcdep.2005.01.008

58. Gorey C, Kroon E, Runia N, Bornovalova M, Cousijn J. Direct Effects of Cannabis Intoxication on Motivations for Softer and Harder Drug Use: An Experimental Approach to the Gateway Hypothesis. Cannabis Cannabinoid Res. Jun 2024;9(3):e830–e838. doi:10.1089/can.2022.0157

59. Vandrey RG, Budney AJ, Hughes JR, Liguori A. A within-subject comparison of withdrawal symptoms during abstinence from cannabis, tobacco, and both substances. Drug Alcohol Depend. Jan 1 2008;92(1-3):48–54. doi:10.1016/j.drugalcdep.2007.06.010

60. Haney M, Bedi G, Cooper ZD, et al. Predictors of marijuana relapse in the human laboratory: robust impact of tobacco cigarette smoking status. Biol Psychiatry. Feb 1 2013;73(3):242–8. doi:10.1016/j.biopsych.2012.07.028

61. Herrmann ES, Cooper ZD, Bedi G, et al. Varenicline and nabilone in tobacco and cannabis co-users: effects on tobacco abstinence, withdrawal and a laboratory model of cannabis relapse. Addict Biol. Jul 2019;24(4):765–776. doi:10.1111/adb.12664

62. Parker MA, Streck JM, Bergeria CL, et al. Reduced Nicotine Content Cigarettes and Cannabis Use in Vulnerable Populations. Tob Regul Sci. Sep 2018;4(5):84–91. doi:10.18001/trs.4.5.8

63. West R, Evins AE, Benowitz NL, et al. Factors associated with the efficacy of smoking cessation treatments and predictors of smoking abstinence in EAGLES. Addiction. Aug 2018;113(8):1507–1516. doi:10.1111/add.14208

64. Constantin J, Jayawardhana J. Cigarette and cannabis use and co-use among U.S. adults: An examination of prevalence and trends during 2015-2023. Addict Behav. Jan 2026;172:108521. doi:10.1016/j.addbeh.2025.108521

65. Rubenstein D, McClernon FJ, Pacek LR. Trends in cannabis and tobacco co-use in the United States, 2002-2021. Addict Behav. Nov 2024;158:108129. doi:10.1016/j.addbeh.2024.108129

66. Panlilio LV, Zanettini C, Barnes C, Solinas M, Goldberg SR. Prior exposure to THC increases the addictive effects of nicotine in rats. Neuropsychopharmacology. Jun 2013;38(7):1198–208. doi:10.1038/npp.2013.16

67. Yeap ZJS, Marsault J, George TP, Mizrahi R, Rabin RA. Does tobacco dependence worsen cannabis withdrawal in people with and without schizophrenia-spectrum disorders? Am J Addict. Jul 2023;32(4):367–375. doi:10.1111/ajad.13394

68. Peng J, Fan M, An C, Ni F, Huang W, Luo J. A narrative review of molecular mechanism and therapeutic effect of cannabidiol (CBD). Basic Clin Pharmacol Toxicol. Apr 2022;130(4):439–456. doi:10.1111/bcpt.13710

69. Laprairie RB, Bagher AM, Kelly ME, Denovan-Wright EM. Cannabidiol is a negative allosteric modulator of the cannabinoid CB1 receptor. Br J Pharmacol. Oct 2015;172(20):4790–805. doi:10.1111/bph.13250

70. Resstel LB, Tavares RF, Lisboa SF, Joca SR, Corrêa FM, Guimarães FS. 5-HT1A receptors are involved in the cannabidiol-induced attenuation of behavioural and cardiovascular responses to acute restraint stress in rats. Br J Pharmacol. Jan 2009;156(1):181–8. doi:10.1111/j.1476-5381.2008.00046.x

71. Marinelli S, Pascucci T, Bernardi G, Puglisi-Allegra S, Mercuri NB. Activation of TRPV1 in the VTA excites dopaminergic neurons and increases chemical- and noxious-induced dopamine release in the nucleus accumbens. Neuropsychopharmacology. May 2005;30(5):864–70. doi:10.1038/sj.npp.1300615

72. Bisogno T, Hanus L, De Petrocellis L, et al. Molecular targets for cannabidiol and its synthetic analogues: effect on vanilloid VR1 receptors and on the cellular uptake and enzymatic hydrolysis of anandamide. Br J Pharmacol. Oct 2001;134(4):845–52. doi:10.1038/sj.bjp.0704327

73. Leweke FM, Piomelli D, Pahlisch F, et al. Cannabidiol enhances anandamide signaling and alleviates psychotic symptoms of schizophrenia. Transl Psychiatry. Mar 20 2012;2(3):e94. doi:10.1038/tp.2012.15

74. Nasrin S, Coates S, Bardhi K, Watson C, Muscat JE, Lazarus P. Inhibition of Nicotine Metabolism by Cannabidiol (CBD) and 7-Hydroxycannabidiol (7-OH-CBD). Chem Res Toxicol. Feb 20 2023;36(2):177–187. doi:10.1021/acs.chemrestox.2c00259

75. Hindocha C, Freeman TP, Grabski M, et al. Cannabidiol reverses attentional bias to cigarette cues in a human experimental model of tobacco withdrawal. Addiction. May 1 2018;113(9):1696–705. doi:10.1111/add.14243

76. Hindocha C, Freeman TP, Xia JX, Shaban NDC, Curran HV. Acute memory and psychotomimetic effects of cannabis and tobacco both ’joint’ and individually: a placebo-controlled trial. Psychol Med. Nov 2017;47(15):2708–2719. doi:10.1017/s0033291717001222

77. Hindocha C, Lawn W, Freeman TP, Curran HV. Individual and combined effects of cannabis and tobacco on drug reward processing in non-dependent users. Psychopharmacology (Berl). Nov 2017;234(21):3153–3163. doi:10.1007/s00213-017-4698-2

78. al’Absi M, DeAngelis BN, Nakajima M, et al. Biobehavioral and affective stress responses during nicotine withdrawal: Influence of regular cannabis co-use. Psychopharmacology (Berl). Feb 2024;241(2):253–262. doi:10.1007/s00213-023-06481-w

