## Supplementary Material for "Cannabis Co-Use and Endocannabinoid System Modulation in Tobacco Use Disorder: A Translational Systematic Review and Meta-Analysis"

### Supplementary Table S1. Database Search Strategies

Searches were conducted on December 13, 2024 and updated on January 15, 2026. The search strategy was structured around two complementary questions: (Q1) the effect of cannabinoids on tobacco/nicotine cessation, reduction, or dependence; and (Q2) the relationship between cannabinoid exposure and nicotine dependence outcomes in experimental studies. The strategies below present the search syntax for each database without result counts.

#### Ovid MEDLINE

| **#** | **Search Terms** |
| --- | --- |
| 1 | *cannabidiol/ or *cannabis/ or *marijuana smoking/ or (cannab* or marijuana or marihuana).tw,kw. |
| 2 | smoking cessation/ or "tobacco use cessation"/ or ((tobacco or nicotine or cig* or ecig* or smoker* or smoking) adj7 (cessation or quit or discontinuation or discontinue or abstain or abstinence)).tw,kw. |
| 3 | smoking reduction/ or ((tobacco or nicotine or cig* or ecig* or smoker* or smoking) adj7 (reduction or reduce or replace or replacement)).tw,kw. |
| 4 | "Tobacco Use Disorder"/ or ((tobacco or nicotine or cig* or ecig* or smoker* or smoking) adj (addict* or disorder* or misuse or abuse or disorder* or dependence)).tw,kw. |
| 5 | 1 and 2 |
| 6 | 1 and 3 |
| 7 | 1 and 4 |
| 8 | 5 or 6 or 7 |
| 9 | exp "tobacco use disorder"/ or ((tobacco or nicotine or cigarett*) adj4 smoking).tw,kf. |
| 10 | (random* or trial* or experiment* or Prospect* or blind*).mp. |
| 11 | 1 and 9 and 10 |
| 12 | 8 or 11 |

#### Embase

| **#** | **Search Terms** |
| --- | --- |
| 1 | *Cannabidiol/ or *cannabis/ or *cannabis smoking/ or (cannab* or marijuana or marihuana).tw,kw. |
| 2 | *Smoking cessation/ or ((tobacco or nicotine or cig* or ecig* or smoker* or smoking) adj7 (cessation or quit or discontinuation or discontinue or abstain or abstinence)).tw,kw. |
| 3 | *Smoking reduction/ or ((tobacco or nicotine or cig* or ecig* or smoker* or smoking) adj7 (reduction or reduce or replace or replacement or decrease)).tw,kw. |
| 4 | *Tobacco dependence/ or ((tobacco or nicotine or cig* or ecig* or smoker* or smoking) adj (addict* or disorder* or misuse or abuse or disorder* or dependenc*)).tw,kw. |
| 5 | 1 and 2 |
| 6 | 1 and 3 |
| 7 | 1 and 4 |
| 8 | 5 or 6 or 7 |
| 9 | *Tobacco dependence/ or ((tobacco or nicotine or cigarett*) adj4 smoking).mp. |
| 10 | (random* or trial* or experiment* or Prospect* or blind*).mp. |
| 11 | 1 and 9 and 10 |
| 12 | 8 or 11 |
| 13 | limit 12 to (conference abstracts or "preprints (unpublished, non-peer reviewed)") |
| 14 | 12 not 13 |

#### APA PsycInfo

| **#** | **Search Terms** |
| --- | --- |
| 1 | Cannabidiol/ or *cannabis/ or "cannabis use"/ or (cannab* or marijuana or marihuana).tw. |
| 2 | Smoking cessation/ or ((tobacco or nicotine or cig* or ecig* or smoker* or smoking) adj7 (cessation or quit or discontinuation or discontinue or abstain or abstinence)).tw. |
| 3 | ((tobacco or nicotine or cig* or ecig* or smoker* or smoking) adj7 (reduction or reduce or replace or replacement or decrease)).tw. |
| 4 | "Tobacco Use Disorder"/ or ((tobacco or nicotine or cig* or ecig* or smoker* or smoking) adj (addict* or disorder* or misuse or abuse or disorder* or dependenc*)).tw. |
| 5 | 1 and 2 |
| 6 | 1 and 3 |
| 7 | 1 and 4 |
| 8 | 5 or 6 or 7 |
| 9 | exp cannabinoids/ or (cannab* or tetrahydrocannabinol or marijuana).tw. |
| 10 | "Tobacco Use Disorder"/ or ((tobacco or nicotine or cigarett*) adj4 smoking).mp. |
| 11 | (random* or trial* or experiment* or Prospect* or blind*).mp. |
| 12 | 9 and 10 and 11 |
| 13 | 8 or 12 |

#### Web of Science Core Collection (Clarivate)

| **#** | **Search Terms** |
| --- | --- |
| 1 | TI=(cannab* OR marijuana OR marihuana) or AB=(cannab* OR marijuana OR marihuana) |
| 2 | TI=((tobacco or nicotine or cig* or ecig* or smoker* or smoking) NEAR/4 (cessation or quit or discontinuation or discontinue or abstain or abstinence)) OR AB=((tobacco or nicotine or cig* or ecig* or smoker* or smoking) NEAR/4 (cessation or quit or discontinuation or discontinue or abstain or abstinence)) |
| 3 | TI=((tobacco or nicotine or cig* or ecig* or smoker* or smoking) NEAR/4 (reduction or reduce or replace or replacement or decrease)) OR AB=((tobacco or nicotine or cig* or ecig* or smoker* or smoking) NEAR/4 (reduction or reduce or replace or replacement or decrease)) |
| 4 | TI=((tobacco or nicotine or cig* or ecig* or smoker* or smoking) NEAR/4 (addict* or disorder* or misuse or abuse or disorder* or dependenc*)) OR AB=((tobacco or nicotine or cig* or ecig* or smoker* or smoking) NEAR/4 (addict* or disorder* or misuse or abuse or disorder* or dependenc*)) |
| 5 | #1 and (#2 or #3 or #4) |
| 6 | TI=((tobacco or nicotine or cigarett*) NEAR/4 (smoking or disorder*)) or AB=((tobacco or nicotine or cigarett*) NEAR/4 (smoking or disorder*)) |
| 7 | TI=(random* or trial* or experiment* or Prospect* or blind*) or AB=(random* or trial* or experiment* or Prospect* or blind*) |
| 8 | #1 AND #6 AND #7 |
| 9 | #5 or #8 |
| 10 | #5 or #8 and Meeting Abstract or Editorial Material or Book Chapters (Exclude – Document Types) |

**Supplementary Table S2.** Characteristics of Observational Studies Examining the Association Between Cannabis Co-Use and Tobacco/Nicotine Cessation Outcomes (k = 18). Cannabis use was assessed as any use (yes/no) across all studies; none provided stratification by quantity or frequency of cannabis use.

| **Study** | **N Total** **(CU / Non-CU)** | **Target** **Product** | **Intervention / Setting** | **Cessation Outcome** **& Verification** | **Cannabis Use Assessment** | **FU** | **OR (95% CI)** | **p** | **Key Covariates** |
| --- | --- | --- | --- | --- | --- | --- | --- | --- | --- |
| Allagbe 2025 | 94,827 (10,239 / 84,588) | Combustible cigarettes | SCS (CBT + pharmacotherapy) | 1-month CA; bio-verified (CO < 5 ppm) | Self-report, past 30-day use; route NR | 4 mo | Diabetic: 0.67 (0.50–0.90) Non-diabetic: 0.71 (0.68–0.75) | 0.008; <0.001 | Age, sex, education, employment, CVD, depression, anxiety, nicotine dependence |
| Gilman 2025 | 251 (178 / 73) | E-cigarettes | Pharmacotherapy (varenicline vs. placebo) + behavioral counseling | 7-day PPA; bio-verified (salivary cotinine) | Self-report (TLFB) + CUDIT-R; route NR | 3 mo | 1.14 (0.51–2.57) | 0.20 | Age, sex, alcohol use, treatment group |
| Graham 2025 | 2,845 (1,840 / 1,005) | E-cigarettes | Digital/remote (“This is Quitting” text-message program) | Dual abstinence (nicotine + cannabis); self-report | Self-report, past 30-day use; route NR | 7 mo | Adolescents: 0.70 (0.52–0.94) Young adults: 0.27 (0.20–0.35) | 0.019; <0.001 | Unadjusted |
| Lambart 2024 | 500 (75 / 425) | Combustible cigarettes | Pharmacotherapy (varenicline vs. placebo) + culturally relevant counseling | 7-day PPA; bio-verified (salivary cotinine ≤15 ng/mL) | Self-report (blunt use) | 3 mo | 0.84 (0.38–1.87) | 0.67 | Treatment assignment |
| Voci 2024 | 83,206 (22,401 / 58,284) | Combustible cigarettes | Pharmacotherapy (NRT + brief counseling) | 7-day PPA; self-report | Self-report, past 30-day use; route NR | 6 mo | 0.93 (0.89–0.97) | 0.001 | Age, gender, employment, income, quit attempts, CPD, TTFC, importance, confidence, hazardous alcohol, opioid use, rurality, treatment setting, psychiatric/medical diagnoses |
| Ozga 2023 | 374 (39 / 176) | Combustible cigarettes | Digital/remote (web-based) + NRT patches | 7-day PPA; bio-verified (CO < 10 ppm; salivary cotinine if needed) | NR | 6 mo | 0.22 (0.03–0.90) | NR | Study site, intervention condition, race, ethnicity, housing stability, smoking partner, other tobacco use |
| Goodwin 2022 | 3,396 (283 / 3,113) | Combustible cigarettes | Digital/remote (quitline + NRT screening) | 30-day PPA; self-report | Self-report, any cannabis form (CBD-only excluded) | 7 mo | 0.68 (0.44–1.05) | 0.08 | Age, sex, race/ethnicity, education |
| McClure 2021 | 1,390 (441 / 949) | Combustible cigarettes | Digital/remote (quitline + web-based program) | 7-day PPA; self-report | NR | 6 mo | 0.98 (0.59–1.62) | 0.926 | None reported |
| Le Faou 2020 | 1,624 (176 / 1,448) | Combustible cigarettes | Pharmacotherapy (NRT, varenicline) + behavioral support | 28-day CA; bio-verified (CO < 10 ppm) | Self-report, past month (yes/no); route NR | 1 mo | 0.61 (0.41–0.88) | <0.05 | Gender, age, education, medical background, medications, quit history, CPD, FTND, self-efficacy, alcohol, pharmacotherapy type, follow-up visits |
| McClure 2020 | 157 (107 / 50) | Combustible cigarettes | Pharmacotherapy (varenicline vs. placebo) + brief counseling | Weekly PPA; self-report | Self-report (past 30-day, days used) + urine THC; route NR | 3 mo | 0.40 (0.22–0.85) | 0.021 | Treatment assignment, baseline CPD, gender, study visit |
| Rogers 2020 | 207 (82 / 125) | Combustible cigarettes | Pharmacotherapy (NRT patch) + CBT-based cessation program | 7-day PPA; self-report (MLM) | Urine THC-COOH + self-report, past 30-day use; route NR | 3 mo | 0.40 (0.25–0.63) | <0.001 | Age, gender, baseline FTCD, treatment condition |
| Voci 2020 | 35,246 (1,066 / 28,061) | Combustible cigarettes | Pharmacotherapy (NRT, personalized) + brief counseling | 30-day PPA; self-report | Self-report, past 30-day recreational/medical use; route NR | 6 mo | 0.86 (0.78–0.95) | 0.002 | Age, sex, income, alcohol, opioid use, chronic pain, psychiatric diagnoses, NRT weeks |
| Vogel 2018 | 500 (254 / 246) | Combustible cigarettes | Digital/remote (Facebook-based TSP) | 7-day PPA; self-report | Self-report, past 30-day use (implied); route NR | 3–12 mo | 0.56 (0.35–0.90) | 0.017 | Treatment condition, stage of change, CPD, sex, alcohol use, age of smoking onset |
| Streck 2017 | 1,357 (415 / 942) | Combustible cigarettes | Pharmacotherapy + digital/remote (IVR calls, quitline referral) | 7-day PPA; bio-verified (cotinine < 10 ng/mL or CO < 9 ppm) | Self-report (past year) or ICD-9 diagnosis; route NR | 6 mo | 0.77 (0.51–1.14) | 0.191 | Study arm, nicotine dependence, CPD, age, gender, race, marital status, education, discharge diagnosis, QoL, home smoking policy, quit plan, heavy alcohol, marijuana use |
| Rabin 2016 | 1,226 (220 / 1,006) | Combustible cigarettes | Pharmacotherapy (varenicline, bupropion, NRT, or placebo) + behavioral counseling | 7-day PPA; bio-verified (CO < 10 ppm) | Self-report, past month use; route NR | 3 mo | 0.80 (0.52–1.25) | 0.33 | Age, gender, race, education, CPD, FTND, home smoking policy, partner smoking, treatment assignment |
| Metrik 2011 | 208 (29 / 179) | Combustible cigarettes | Pharmacotherapy (NRT patch) + individual cessation counseling | 7-day PPA; bio-verified (CO + cotinine) | Self-report (TLFB, 8-week window); route NR | 6.5 mo | 1.13 (0.63–2.04) | 0.69 | Gender, age, FTND, treatment condition |
| Okoli 2011 | 239 (~44 / NR) | Combustible cigarettes | Pharmacotherapy (NRT, oral meds) + group behavioral counseling | 7-day PPA; bio-verified (CO ≤8 ppm) | NR | 6.5 mo | 0.17 (0.03–1.28) | NR | Primary substance use, psychiatric diagnoses, evidence-based modality use, FTND, baseline CO, program visits |
| Gourlay 1994 | 1,481 (142 / 1,339) | Combustible cigarettes | Pharmacotherapy (nicotine patch) + brief counseling | 28-day sustained abstinence; bio-verified (CO < 8 ppm) | NR (described as "marijuana smoking") | 6.5 mo | 0.40 (0.20–0.80) | 0.005 | Age, sex, education, motivation, CPD, depression, alcohol consumption |

***Abbreviations:*** *CU, cannabis users; Non-CU, non-cannabis users; CA, continued abstinence; PPA, point prevalence abstinence; CO, carbon monoxide; CPD, cigarettes per day; FTND, Fagerström Test for Nicotine Dependence; FTCD, Fagerström Test for Cigarette Dependence; NRT, nicotine replacement therapy; NR, not reported; OAT, opioid agonist therapy; CBT, cognitive-behavioral therapy; SCS, smoking cessation service; QoL, quality of life; TTFC, time to first cigarette; CVD, cardiovascular disease; IVR, interactive voice response; TSP, Tobacco Status Project; MLM, multilevel model; TLFB, Timeline Followback; CUDIT-R, Cannabis Use Disorder Identification Test-Revised; THC-COOH, 11-nor-9-carboxy-Δ9-tetrahydrocannabinol; FU, follow-up; mo, months.*

***Notes:*** *All odds ratios are expressed as OR for abstinence (cannabis users vs. non-users), with values < 1 indicating lower odds of cessation among cannabis users. For Allagbe 2025, results are presented separately for diabetic and non-diabetic subsamples. For Graham 2025, results are presented separately for adolescent and young adult subsamples; outcome was dual abstinence (nicotine and cannabis); crude ORs are reported (unadjusted). For Okoli 2011, the number of cannabis users was estimated from Table 1 of the original publication (17.0% of N = 258).*

### Supplementary Figure S1. Funnel Plot for Publication Bias Assessment

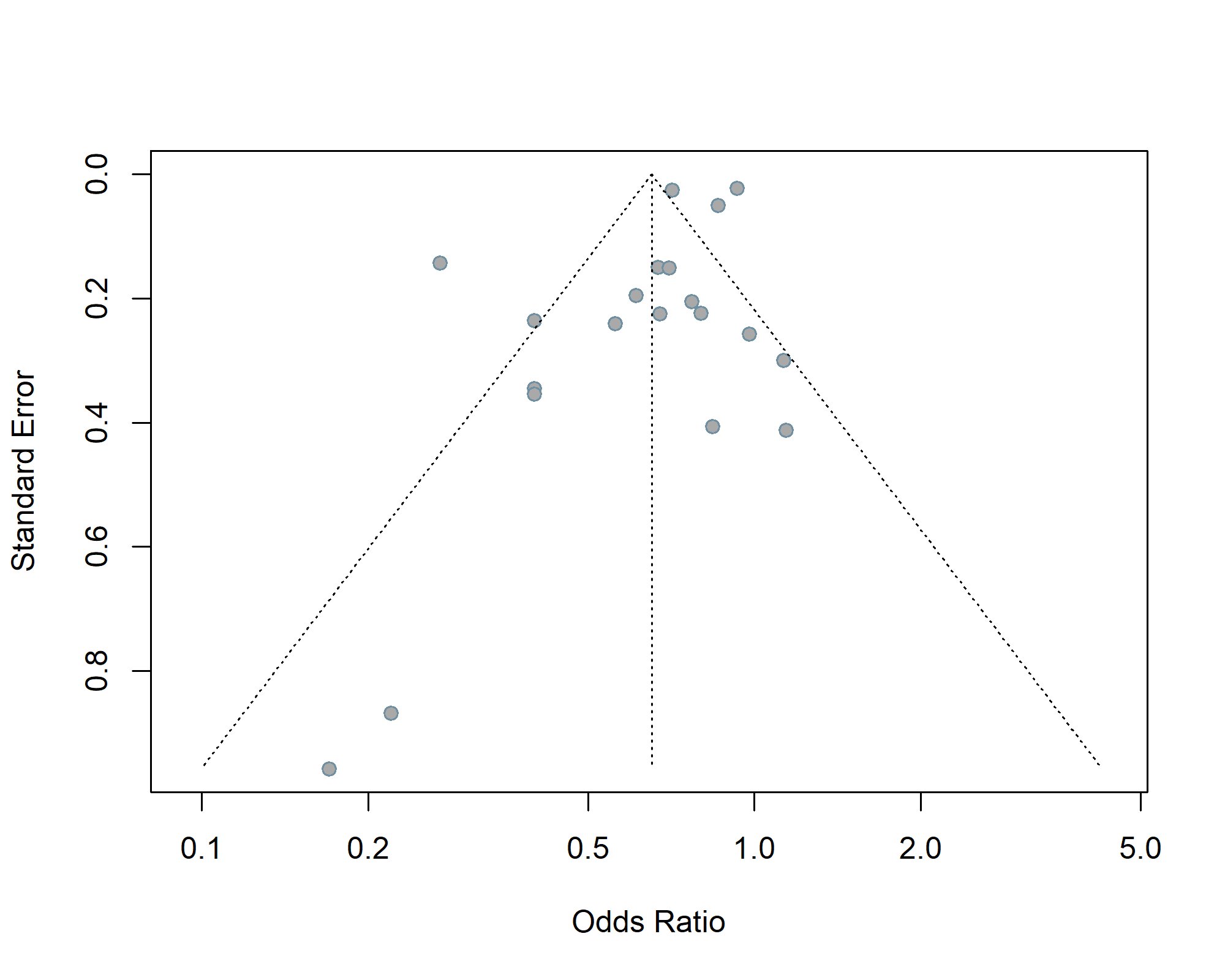

**Supplementary Figure S1.** Funnel plot assessing publication bias across studies included in the meta-analysis of the effects of cannabinoids on tobacco/nicotine cessation outcomes. Each point represents an individual study, plotted as the odds ratio (x-axis, log scale) against the standard error (y-axis, inverted). The dotted lines represent the pseudo 95% confidence limits around the pooled estimate (vertical dotted line). Visual inspection of the funnel plot did not reveal marked asymmetry, and neither Egger’s regression test (p = 0.071) nor Begg’s rank correlation test (p = 0.948) indicated significant publication bias.

### Supplementary Table S3. Risk of Bias Assessment for Randomized Controlled Trials (RoB 2)

| **Study (Author, Year)** | **D1: Randomization Process** | **D2: Deviations from Intended Interventions** | **D3: Missing Outcome Data** | **D4: Measurement of the Outcome** | **D5: Selection of the Reported Result** | **Overall Risk of Bias** |
| --- | --- | --- | --- | --- | --- | --- |
| **Morgan et al. (2013)** | Some concerns | Low | Low | Low | Low | **Some concerns** |
| **Hindocha et al. (2018) [Addiction]** | Low | Low | Low | Low | Low | **Low** |
| **Hindocha et al. (2018) [Sci Rep]** | Low | Low | Low | Low | Low | **Low** |
| **Gournay et al. (2024)** | High | High | Low | Some concerns | Some concerns | **High** |
| **Robinson et al. (2017)** | Low | Low | Some concerns | Low | Some concerns | **Some concerns** |
| **Morrison et al. (2010)** | Low | Low | Low | Low | Low | **Low** |
| **Tonstad & Aubin (2012)** | Low | Low | Low | Low | Low | **Low** |
| **Kelly et al. (1990)** | Low | Low | Low | Low | Low | **Low** |
| **Nemeth-Coslett et al. (1986)** | Low | Low | Low | Low | Low | **Low** |
| **Peters et al. (2021)** | Some concerns | Low | Low | Low | Low | **Some concerns** |
| **Gorey et al. (2024)** | Some concerns | Low | Low | Low | Low | **Some concerns** |
| **Herrmann et al. (2019)** | Low | Low | Some concerns | Low | Low | **Some concerns** |
| **Hindocha et al. (2017) [Psych Med]** | Low | Low | Low | Low | Low | **Low** |
| **Hindocha et al. (2017) [Psychopharm]** | Low | Low | Low | Low | Low | **Low** |
| **Penetar et al. (2005)** | Low | Low | Low | Low | Low | **Low** |
| **Vandrey et al. (2008)** | Some concerns | Low | Some concerns | Low | Some concerns | **Some concerns** |
| **al'Absi et al. (2024)** | Some concerns | Low | Low | Low | Low | **Some concerns** |
| **Haney et al. (2013)** | Some concerns | Low | Some concerns | Low | Some concerns | **Some concerns** |
| **Parker et al. (2018)** | Some concerns | Low | Low | Low | Low | **Some concerns** |

*Color coding: Green = Low risk; Amber = Some concerns; Red = High risk.*

### Supplementary Table S4. Risk of Bias Assessment for Observational Studies (ROBINS-I)

| **Study (Author, Year)** | **D1: Confounding** | **D2: Selection of Participants** | **D3: Classification of Interventions** | **D4: Deviations from Intended Interventions** | **D5: Missing Data** | **D6: Measurement of Outcomes** | **D7: Selection of the Reported Result** | **Overall Risk of Bias** |
| --- | --- | --- | --- | --- | --- | --- | --- | --- |
| **Okoli (2011)** | Serious | Moderate | Moderate | Low | Serious | Low | Moderate | **Serious** |
| **Ozga (2023)** | Low | Low | Low | Low | Low | Low | Low | **Low** |
| **Graham (2025)** | Serious | Low | Serious | Low | Moderate | Serious | Moderate | **Serious** |
| **Rogers (2020)** | Moderate | Moderate | Moderate | Low | Moderate | Serious | Low | **Serious** |
| **McClure (2020)** | Serious | Moderate | Moderate | Low | Moderate | Serious | Moderate | **Serious** |
| **Gourlay (1994)** | Moderate | Moderate | Serious | Low | Moderate | Low | Low | **Serious** |
| **Vogel (2018)** | Serious | Moderate | Moderate | Low | Moderate | Serious | Moderate | **Serious** |
| **LeFaou (2020)** | Moderate | Low | Moderate | Low | Serious | Low | Low | **Serious** |
| **Allagbe (2025)** | Moderate | Low | Moderate | Low | Moderate | Low | Low | **Moderate** |
| **Goodwin (2022)** | Moderate | Moderate | Moderate | Low | Critical | Serious | Moderate | **Serious** |
| **Streck (2017)** | Moderate | Low | Moderate | Low | Moderate | Low | Low | **Moderate** |
| **Rabin (2016)** | Moderate | Moderate | Low | Low | Moderate | Low | Low | **Moderate** |
| **Lambart (2024)** | Moderate | Moderate | Moderate | Low | Moderate | Low | Low | **Moderate** |
| **Voci (2020)** | Moderate | Low | Moderate | Low | Serious | Serious | Low | **Serious** |
| **Voci (2024)** | Moderate | Low | Moderate | Low | Moderate | Serious | Low | **Serious** |
| **McClure (2021)** | Moderate | Moderate | Serious | Low | Moderate | Serious | Low | **Serious** |
| **Metrik (2011)** | Moderate | Serious | Moderate | Low | Moderate | Low | Low | **Serious** |
| **Gilman (2025)** | Moderate | Low | Moderate | Low | Moderate | Low | Low | **Moderate** |

*Color coding: Green = Low risk; Amber = Moderate risk; Red = Serious risk; Dark Red = Critical risk; Gray = No information.*
